# iCVS - Inferring Cardio-Vascular hidden States from physiological signals available at the bedside

**DOI:** 10.1101/2022.12.31.22284089

**Authors:** Neta Ravid Tannenbaum, Omer Gottesman, Azadeh Assadi, Mjaye Mazwi, Uri Shalit, Danny Eytan

## Abstract

Intensive care medicine is complex and resource-demanding. A critical and common challenge lies in inferring the underlying physiological state of a patient from partially observed data. Specifically for the cardiovascular system, clinicians use observables such as heart rate, arterial and venous blood pressures, as well as findings from the physical examination and ancillary tests to formulate a mental model and estimate hidden variables such as cardiac output, vascular resistance, filling pressures and volumes, and autonomic tone. Then, they use this mental model to derive the causes for instability and choose appropriate interventions. Not only this is a very hard problem due to the nature of the signals, but it also requires expertise and a clinician’s ongoing presence at the bedside. Clinical decision support tools based on mechanistic dynamical models offer an appealing solution due to their inherent explainability, corollaries to the clinical mental process, and predictive power. With a translational motivation in mind, we developed iCVS: a simple, with high explanatory power, dynamical mechanistic model to infer hidden cardiovascular states. Full model estimation requires no prior assumptions on physiological parameters except age and weight, and the only inputs are arterial and venous pressure waveforms. iCVS also considers autonomic and non-autonomic modulations. To gain more information without increasing model complexity, both slow and fast timescales of the blood pressure traces are exploited, while the main inference and dynamic evolution are at the longer, clinically relevant, timescale of minutes. iCVS is designed to allow bedside deployment at pediatric and adult intensive care units and for retrospective investigation of cardiovascular mechanisms underlying instability. In this paper, we describe iCVS and inference system in detail, and using a dataset of critically-ill children, we demonstrate its use and power to identify bleeding, distributive states, and cardiac dysfunction, in isolation and in combination.

**Author summary:** A common challenge clinicians face across different disciplines is estimating the hidden physiological state of a patient based on partially observed data. Here we describe iCVS (inferring Cardio-Vascular States): a dynamical mechanistic model of the cardiovascular system. We developed iCVS with a translational goal in mind, showing high explanatory power, its inference relies only on routinely available signals, and enables the identification of various clinically important shock states. We demonstrate the use of the model on a dataset that was collected in a pediatric intensive care unit.

## Introduction

### Clinical challenge

Intensive care medicine is complex, resource demanding and expensive. High-stakes decisions need to be made rapidly based on the evolving clinical state of the patient and the interpretation by clinicians of continuous physiological and ancillary data. The patient’s cardiovascular state may fluctuate over minutes with rapid transitions due to external perturbations, internal failures of the physiological sub-components, or their control. Moreover, these patients are very diverse in their physiological and disease processes. Inferring the underlying hidden physiological state of a patient from observed data is a critical problem that is encountered by clinicians across different disciplines daily. While the modality of the available data and the frequency of sampling may vary between different clinical scenarios, in almost all cases clinicians must rely on partial observations that are a peripheral reflection of the internal, hidden states that are of importance. Specifically, clinicians use observable measurements of heart rate, arterial and venous blood pressures as well as findings from the physical examination (capillary refill, extremity temperature, etc..) to estimate internal variables that are not readily observable at the bedside such as cardiac output, systemic vascular resistance, filling pressures and volumes, autonomic tone, and more. As the physiological signals are often noisy and ambiguous, creating these mental models is a notoriously hard task that requires clinical expertise. Moreover, since the patient’s state fluctuates frequently, repeat assessments by clinicians are required. The clinicians then use this mental model of the patient’s cardiovascular internal state to derive the causes of instability such as shock type (hypovoloemic, cardiac or distributive for example), and choose appropriate treatments and interventions.

### Modeling Background

The large amount of data available from ICUs motivated researchers from computational disciplines to develop mathematical tools which aim to support decision-making in this area. An appealing approach to the analysis of continuous physiological times series relies on mechanistic modeling, grounded in a biophysical understanding of systems. Mechanistic modeling of the cardiovascular system often requires an understanding of the cardiac cycle as a pump coupled to the arterial and venous conductance systems with associated autonomic nervous control. This understanding is derived from years of experiments analyzing human and animal cardiovascular physiology. Such models are usually based on sets of ordinary differential equations (ODEs). In a sense, they simulate the thought process of the critical-care physician at the bedside trying to estimate the underlying pathophysiology of the patient to tailor care. However, due to the complexity of even the simplest models and the number of unobserved variables and unknown parameters, such mechanistic models were mostly published in the context of well-curated “toy” datasets and in-silico simulations ([1]; [2]; [3]; [4]; [5]; barring a few exceptions. See also the thorough review by Chase et al [4]).

Although simplified mechanistic models of the cardiovascular system show promise, so far, to the best of our knowledge, there are none that have been developed specifically for, and tested on data collected from critically-ill patients at the bedside, nor validated in real-life settings. There are several challenges standing in the way of using mechanistic models at the ICU bedside based on real-time data; these have prevented their deployment so far, and also stood in the way of using them to gain insights into the physiology of critical illness. First, as noted above, mechanistic models require estimating multiple hidden physiological parameters and variables: inverse problems are notoriously difficult, and their solutions are often non-unique. To overcome such difficulties, prior works reduced the number of free parameters by assuming constant values for some of the parameters, often taken from the literature to describe a “typical subject” [2], [5], [6], [7], [8]. However, physiological properties can vary considerably between subjects, and attempting to use a “one size fits all” parameter often fails to properly characterize most patients. An example where this problem is particularly acute is in pediatric critical care units, where patient weight and normative values can vary significantly [9], [10], [11]. Thus, for a model to be useful at the bedside, minimal apriori assumptions regarding the values of hidden parameters must be made. Even so, most published models are still quite complex and require estimation of many hidden variables, and thus rely on sampling of multiple invasive variables that are not readily available outside of a dedicated, specialized laboratory. Many of these models are extremely complex, as they attempt to capture cardiovascular dynamics at second or even sub-second resolution, containing multiple coupled differential equations and hidden parameters rendering them nonidentifiable. On the other hand, limiting model complexity to reduce the number of estimated parameters and variables entails a reduction in the explanatory power of such models. Thus, there is a delicate balance as a result of this trade-off between model complexity and estimability using readily available observations.

Second, the measurement frequency of physiological variables in the clinical world varies from continuous monitoring of vital signs to imaging techniques which are conducted once a day or even less. For example, in the ICU, blood pressure and arterial blood oxygen saturation are continuously monitored, while additional indirect estimation of cardiac function or volumes using tools such as echocardiograms are usually performed infrequently, every several hours to days. Models which rely on low-frequency measurements can be good for chronic conditions such as heart failure [2]. However, to estimate the state of unstable patients, mechanistic models should rely only on measurements with high temporal resolution. Of note, up until recently, [12] even these measurements that are commonly captured by the patient’s bedside monitor were not available for offline analysis or real-time ingestion for model inference.

An additional barrier to adapting cardiovascular mechanistic models to bedside use lies in the need to account for various modes of autonomic nervous control: most previously published work focused on a single, simple feedback loop, neglecting other common autonomic modulations.

### Our approach: iCVS

Our motivation is translational in nature - develop an inference model that can estimate the hidden cardiovascular state, its control, and any causes for instability, in real-time and using only measurements that are readily available at the bedside for critically-ill patients. Such a model can be used both as a clinical support tool at the bedside, and to foster an understanding of the (patho-)physiology of critical illness. We derive a simple model (iCVS), which retains a high explanatory power of the cardio-vascular system including cardio-vascular control, which can capture different clinical scenarios such as bleeding or evolving hypovolemia, cardiogenic shock and distributive shock, in isolation or in combinations. iCVS takes into account the autonomic control of the cardiovascular system, which couples the dynamics of its different components, mathematically imposing coupling constraints on inferred parameters. This reduces the effective dimensionality of the parameter space over which we look for a solution to an inverse problem, allowing us to infer a larger set of unobserved parameters compared to previous models. The estimated parameters enable us to capture individual patients’ traits.

iCVS is designed to allow deployment at the bedside in the pediatric and adult intensive care unit: no prior assumptions regarding the physiological parameters of the patients are required except age and weight, and the only inputs which are required for the estimation process are arterial and venous blood pressures waveforms, which are commonly measured in the ICU. In order to gain more information about the cardiovascular state, without increasing model complexity, we introduce a novel approach that exploits both slow and fast timescales of the blood pressure trace. From the pulse contour shape of the blood pressure waveform, we extract quantities related to the peripheral resistance, stroke volume, and pulse pressure, while the main mathematical model and its inference run using the mean, smoothed, arterial and venous pressures at the longer, clinically-relevant, time scale of minutes. Thus, the main model and its inference focus on processes that evolve over minutes, and are of interest to clinicians, and further allow for a much simpler model than those that capture all the intricacies of each cardiac cycle [2]. All this is done while still capitalizing on information that can be extracted only from the arterial pressure waveform.

In what follows, we describe the iCVS model and inference system in detail and demonstrate its use and power for real-time estimation using a dataset of 10 patients admitted to a pediatric intensive care unit. While our estimation process entails inferring the full iCVS model including 15 hidden parameters, we focus our demonstration on the identification of three clinical shock states, representing a dire, hard clinical challenge, and a testable task. Shock, due to any cause is a state in which the cardiovascular system fails to deliver an adequate supply of oxygen to the tissues. There are several prototypical physiological shock states, each requiring specific interventions, without which there is a high risk for mortality or morbidity. Identifying shock and its causes is a notoriously hard task due to the nature of the observed physiological signals (demonstrated below), requiring continuous vigilance and clinical expertise. Specifically, using iCVS, we map certain hidden parameter ranges to identify (i) ongoing bleeding, (ii) distributive shock, and (iii) cardiogenic shock due to reduced cardiac contractility.

## Results

### iCVS: Model overview

The goal of this work is to derive a new model of the cardiovascular system which can be deployed at the patient’s bedside, requires only routinely acquired measurements as input, and provides an inference of the hidden, internal physiological state of the patient in real-time. The model should be flexible enough to capture the different types of shock and account for the inter-subject variability which characterizes the patients in the pediatric and adult critical care unit. The full details of the model and its development are provided in the Materials and Methods section.

Mechanistic models of the cardiovascular system are developed from first principles, tie together changes in blood pressures and volumes, and usually use the latter as the dependent dynamical variables in the set of ODEs [5]. We derive the model in terms of intravascular pressures since arterial and venous blood pressures are directly measurable (in contrast to blood volume). Briefly, our proposed model contains a one-chamber heart that acts like a pump [5], an arterial component and a venous component, and further assumes the peripheral organs can be lumped as a linear resistor (see Fig 2). In addition and importantly, the model contains three regulatory units which are described below. The two dynamical equations which describe the time-dependent evolution of the intravascular pressures are written as follows (the full derivation can be found in *Materials and methods*):

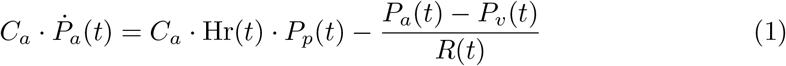

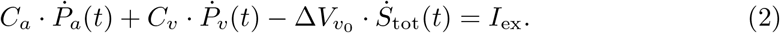

Here *P*_*a*_ and *P*_*v*_ are the mean arterial and venous pressures respectively, Hr is the heart rate, *R* is the systemic vascular resistance and *P*_*p*_ is the pulse pressure. In addition, *C*_*a*_ and *C*_*v*_ are the arterial and venous compliances respectively, Δ*V*_*v*0_ is the difference between minimal and maximal unstressed venous volume, *I*_ex_ is the intravascular volume change, and *S*_tot_ serves as a general marker for the activation of the autonomic nervous system (see below).

Our proposed model contains fifteen hidden parameters (see Table 3), and five time-dependent observables: mean arterial pressure, mean venous pressure, heart rate, peripheral resistance, and pulse pressure (Table 2). We later show how the mechanistic model can be used to estimate the hidden parameters based on the observables, and identify clinical states based on the observed data (Fig 1).

**Fig 1.**
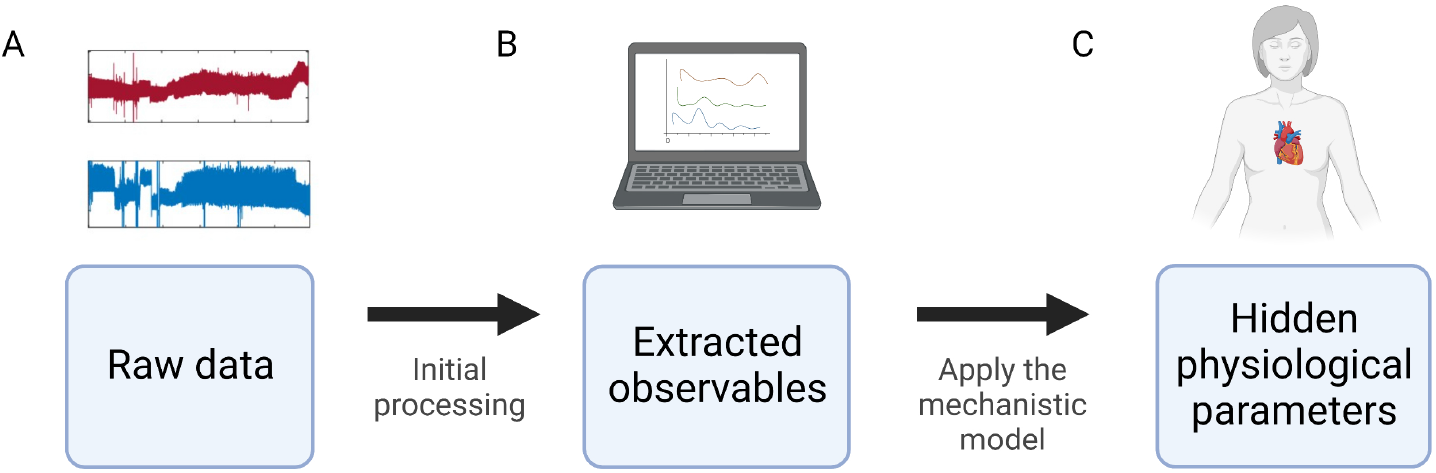
Overview of the estimation process. **A**. Arterial and venous pressure waveforms are recorded via arterial and central venous catheters which connected to pressure transducers. Specifically, in this paper waveforms are sampled from critically-ill children hospitalized in an intensive care unit and recorded at 125Hz. **B**. The raw data is analyzed and five measurements are extracted - the observables. **C**. The observables are used as an input for the full model (iCVS) estimation and using constrained nonlinear optimization we obtain the set of parameter values that best fit observables. *Created with BioRender.com*

**Fig 2.**
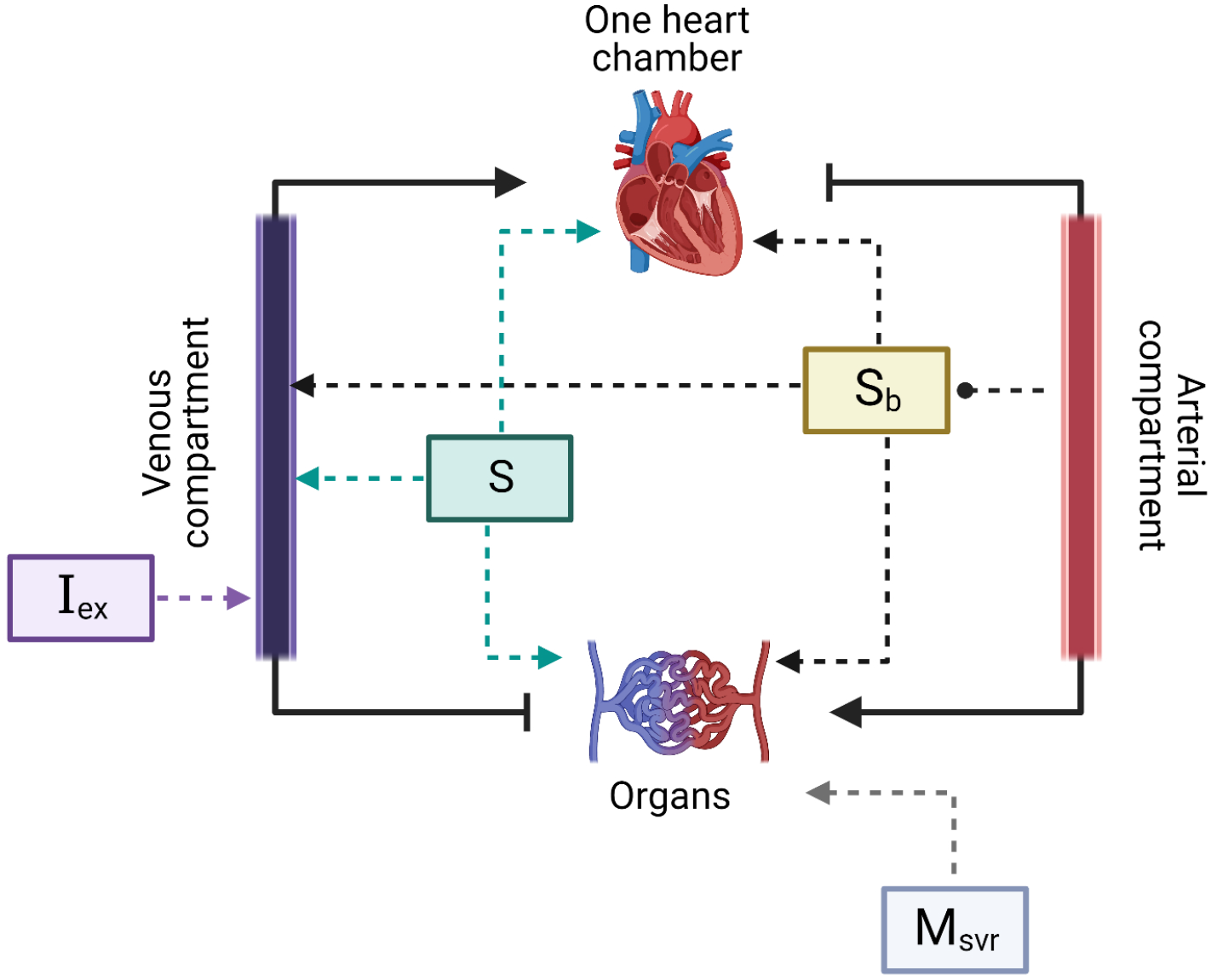
A schematic representation of the cardiovascular model. The model is a serial circuit and contains one heart chamber which acts like a pump, organs, and arterial and venous compartments. Heart rate, contractility, vascular resistance and unstressed venous volume are affected by the baroreflex (*S*_b_) which is determined by the arterial pressure, and by *S* which is an independent component that does not depend on the current physiological state. In addition, intra-vascular volume can change through *I*_ex_ and the vascular resistance is also modulated by *M*_SVR_. *Created with BioRender.com*

### Autonomic regulation of the cardiovascular system in iCVS

In our model, the autonomic nervous system’s activation affects heart rate, cardiac contractility, unstressed venous volume, and peripheral resistance. It consists of two components: (i) The baroreflex (*S*_*b*_) – the internal feedback loop that modulates heart rate, contractility, unstressed venous volume, and the systemic vascular resistance (*R*), meant to keep the mean arterial blood pressure as close to a set-point (*P*_set_) as possible. The baroreflex represents the autonomic component that is dependent on the patient’s current cardiovascular state. (ii) An independent component (*S*), representing the sum of all autonomic modulation which is independent of the cardiovascular state. While the baroreflex *S*_*b*_ describes the activation of different components in response to a reduction in the arterial blood pressure, and thus manifests as negative correlations between the arterial blood pressure and the heart rate (as well as other variables), the independent autonomic component *S* **does not** depend on the arterial blood pressure and thus enables the model to capture different interactions between the blood pressure and heart rate, as well as other cardio-vascular components (Fig 2). For example, this component is expected to rise when sympathetic agonists are given, or when a “central arousal state” is present due to patient agitation or in response to a noxious stimulus (a typical fight or flight response).

### Non-Autonomic regulation of the cardiovascular system in iCVS

In iCVS, in addition to the autonomic control, the systemic vascular resistance is independently modulated by a component denoted by *M*_SVR_ (Fig 2). This component enables the model to describe fluctuations in the peripheral resistance which are uncoupled from the autonomic activation. This can occur for example due to maladaptive vasodilatation that occurs during septic, vasoplegic, or anaphylactic shock, or as a response to intravenous infusion of specific drugs that affect SVR such as Norepinephrine or Vasopressin. Of note, a non-autonomic regulation on the systemic vascular resistance appears also in [7], in the context of metabolic consumption during physical activity. Total blood volume can be changed by an external current *I*_ex_ which can be either positive, as is the case when intravenous fluids are bolused, or negative, such as during bleeding or loss of intravascular volume due to capillary leak or severe dehydration.

### iCVS can simulate different shock states

The interplay between the fifteen parameters of the model can generate different clinical scenarios. In this work we focus on three different shock states which are described by three hidden parameters. Briefly, shock is a failure of the cardiovascular system to supply adequate blood and oxygen to the body tissues. In order to describe the hypovolemic shock state, we use *I*_ex_ (intra-vascular volume change), which is negative in case of intravascular fluid or blood loss. A distributive shock state is equivalent to a negative *M*_SVR_ (non-autonomic vascular resistance modulation). In the next section, we also discuss the cardiogenic shock state which is caused by a reduction in *K* (heart contractility).

Fig 3 presents examples of different shock conditions of artificial neonate patients which are generated by the mechanistic model. The internal parameters of the subjects are set and two of them (*I*_ex_ and *M*_SVR_) are presented (Fig 3A,B,I,J,Q,R). The observable parameters (Fig 3C-G, K-O, S-W) are simulated according to the iCVS model (see methods). The baroreflex, which depends on the current arterial pressure and affects the observables is also presented (Fig 3H,P,X).

**Fig 3.**
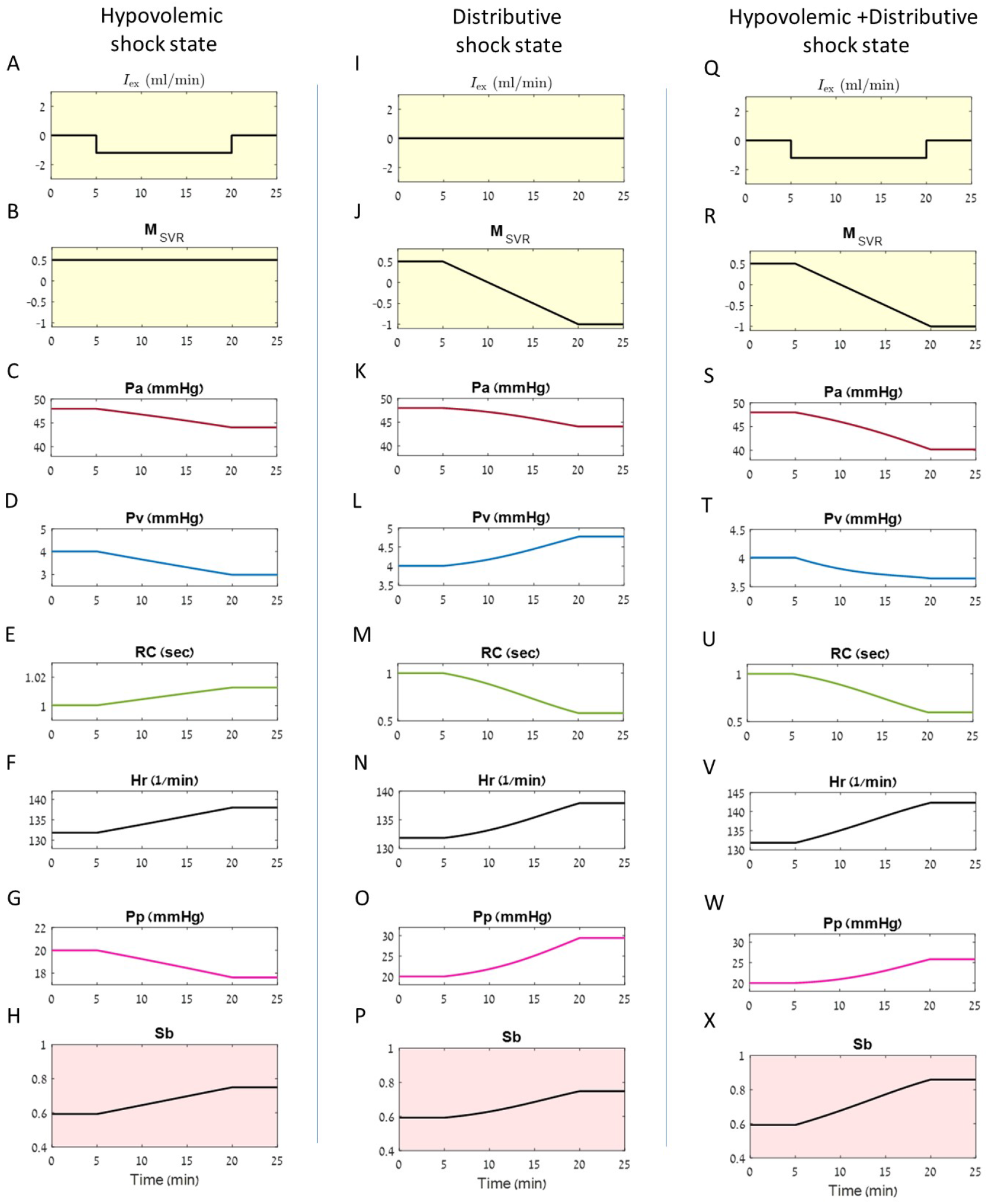
Simulations of the iCVS model for different shock conditions. Each column presents a simulation of one artificial patient with a given shock state: **A-H** - hypovolemic shock state, **I-P** - distributive shock state and **Q-X** - combined hypovolemic and distributive shock state. The hidden parameters are fixed, and the observables are simulated accordingly (see Methods). Two of the hidden parameters are presented for each patient: **A, I, Q** Time dependent intra vascular volume change (*I*_ex_). In **A, Q** liquid withdrawal occurs over a period of 15 minutes. **B, J, R** Time course of *M*_SVR_. In **J, R** a reduction in *M*_SVR_ occurs over a period of 15 minutes. **C-H, K-P, S-X** - the resulting observables: **C, K, S** Arterial pressure (*P*_*a*_), **D, L, T** Venous pressure (*P*_*v*_), **E, M, U** Peripheral resistance multiplied by arterial compliance (*RC*), **F, N, V** Heart rate (Hr), **G, O, W** Pulse pressure (*P*_*p*_). **H, P, X** The time dependent magnitude of the baro-reflex (*S*_*b*_).

Panels A-H in Fig 3 present a simulation of a patient in a hypovolemic shock state, as can happen with ongoing bleeding. The intravascular volume change (*I*_ex_) is negative (Fig 3A), representing a reduction of intravascular volume over time, while the non-autonomic SVR modulation (*M*_SVR_) remains constant (Fig 3B). As expected, we see that the arterial and venous pressures decrease (Fig 3C,D). The heart rate and the vascular resistance reflexively rise (Fig 3E,F) due to baroreflex activation which acts to maintain blood pressure and compensates for the reduction in the intravascular volume (Panel H). Note, that despite the baro-reflex activation that increases the contractility, the pulse pressure (Fig 3)G) decreases due to the change in venous pressure and return (see also Eq 22 in *Material and methods*.

Panels I-P in Fig 3 present a simulation of a patient with a distributive shock state. The parameter *I*_ex_ is zero (Fig 3I), reflecting constant intravascular volume, while *M*_SVR_ decreases (Fig 3J), reflecting a reduction in the peripheral resistance which is not caused by the autonomic nervous system. It is possible to see in Fig 3N that the heart rate increases (due to the baroreflex activation, see panel H), the pulse pressure increases as well (due to the baroreflex and rise in venous pressure), and the peripheral resistance decreases due to the reduction in *M*_SVR_ (Fig 3M).

Panels Q-X in Fig 3 present a simulation of a patient with a combined hypovolemic and distributive shock state. The reduction in the arterial blood pressure(Panel s in Fig 3) is more pronounced than in either shock state separately.

Of note, in addition to simulating shock states, iCVS can emulate common clinical behaviors, such as a fight-or-flight response that is elicited by pain or a noxious stimulus, mediated by autonomic tone modulation (see S1 Fig.).

### Estimation pipeline of the cardiovascular parameters from bedside-acquired blood pressure waveforms

The estimation pipeline is schematically illustrated in Fig. 1 and is described below and in *Material and Methods*. Briefly, the first step includes the initial processing of the raw data and extraction of five observables. In the next step, the observables serve as an input to an optimization function which is based on iCVS and we estimate the hidden parameters and the full model.

#### Extracting the observables from the raw data

As a first step, we use waveforms of venous and arterial blood pressure tracings, acquired at a high frequency that allow extraction of the following five observables: mean arterial pressure, mean venous pressure, heart rate, peripheral resistance, and pulse pressure (see *Material and Methods*). We then use both slow and high-frequency features of the data (Fig 4). From the slow time scale we calculate the mean arterial and venous pressures. From the fast timescale (the scale of a single heartbeat), we extract: heart rate (timing of consecutive beats), peripheral vascular resistance (up to a constant, as detailed in *Material and methods*), and pulse pressure. These quantities, which cannot be derived without the waveforms recorded at high-frequency (a sampling rate which is much higher than the heart rate and can capture the shape of a single beat), are then smoothed and used for model inference at the slower timescale of minutes. The peripheral resistance is extracted using a non-calibrated pulse contour analysis method relying on assumptions of a simplified Windkessel model and rectangular ejection pattern during systole that has been described in detail elsewhere [13, 14].

**Fig 4.**
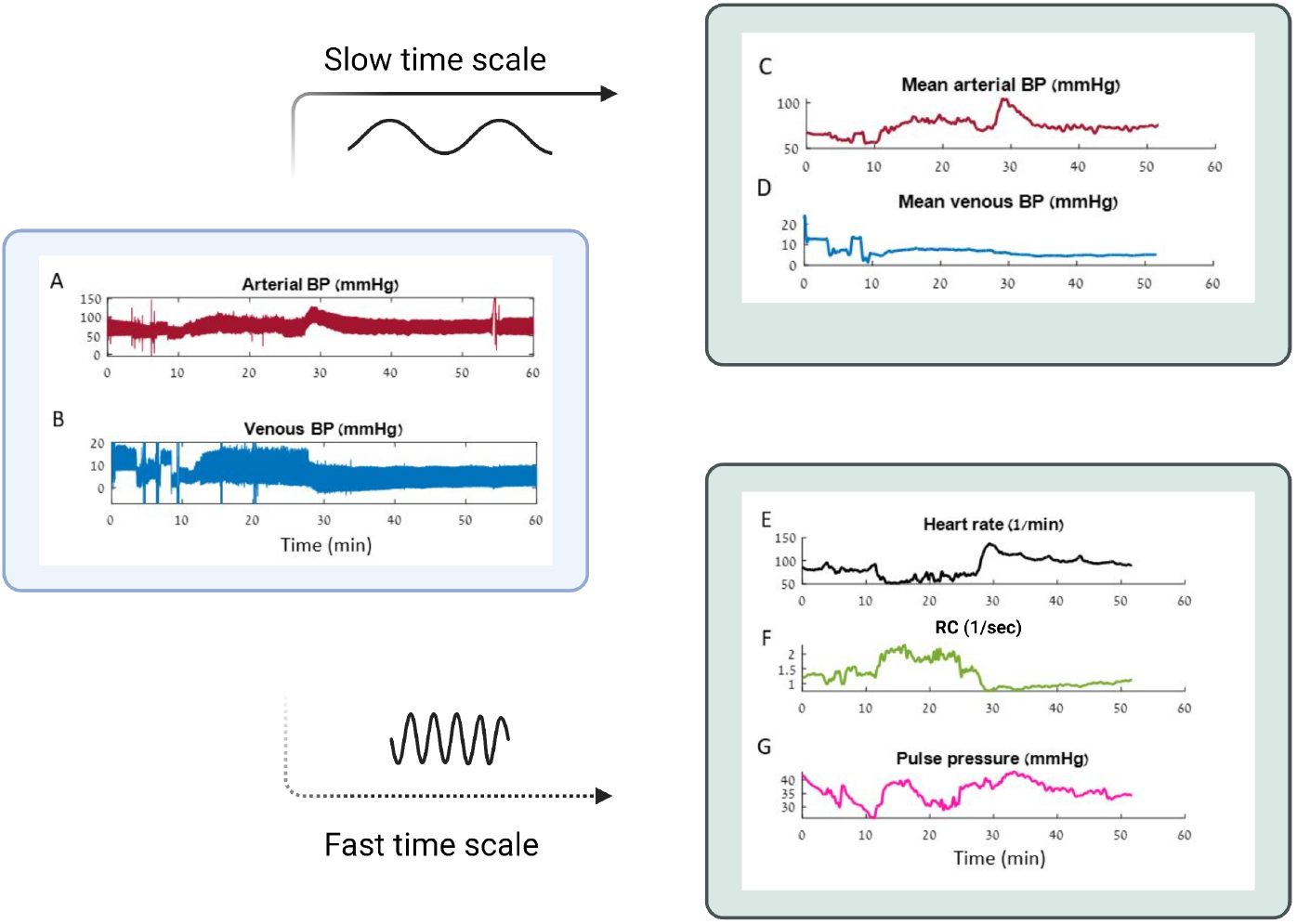
Extraction of slow and fast time scales features from venous and arterial blood pressure recordings. **A**,**B**. An example of the raw data, sampled at 125 Hz frequency by the bedside patient monitor. **A**. Arterial blood pressure, **B**. Venous blood pressure. Note how these real-life data are noisy, fluctuating, and riddled with artifacts related to patient movement and care. **C-G**. Features that are extracted from the raw data. **C**,**D**. Mean arterial and venous pressures (slow time scale features), **E-G**. Heart rate, peripheral resistance and pulse pressure (fast time scale features).

#### Estimating the hidden cardiovascular parameters

The next step in the estimation pipeline is to use the above five observables, as well as information on a patient’s age and weight, to fully estimate the hidden parameters of the iCVS model. Based on the iCVS model, we derive a system of equations that relates the time-dependent measurements to the fifteen hidden physiological parameters (see *Material and methods*). During the estimation procedure, a set of optimal parameter values that yield the best solution for the time-dependent equations is chosen by minimizing a constrained nonlinear multivariate cost function (see more details in *Material and Methods*). For the illustrations shown below, the estimation procedure is independently conducted on data segments of 300 seconds. We chose a segment size of 300 seconds after an empirical search for a window that averaged out noisy and artifactual fast fluctuations, while still capturing changes over clinically-relevant timescales of several minutes.

### Demonstrating iCVS estimation of cardiovascular shock states on real-world data from critically-ill patients

In this section, we illustrate the estimation process for a sample of 10 critically-ill children, hospitalized in an intensive care unit (ICU) at a large, tertiary children’s hospital. These children were admitted to the ICU either for routine observation post-operatively (for the control samples) and were stable hemodynamically, or presented with various shock states, for example, due to massive post-operative bleeding, cardiac dysfunction, infections or other causes, see Table 1 for more information. The dataset which we used to test the model’s performance is detailed in [12]. It contains physiological waveforms recorded from critically-ill children at sampling rates of 125Hz for all invasively-recorded pressures; however, any high-frequency recording that contains the full arterial blood pressure waveform is adequate for the model’s purposes. In addition, the times of all medical interventions, including drug consumption and fluid administration are recorded. Note how these real-life data are noisy, fluctuating and riddled with artifacts related to patient movement and care (see Fig 4A,B).

**Table 1.**
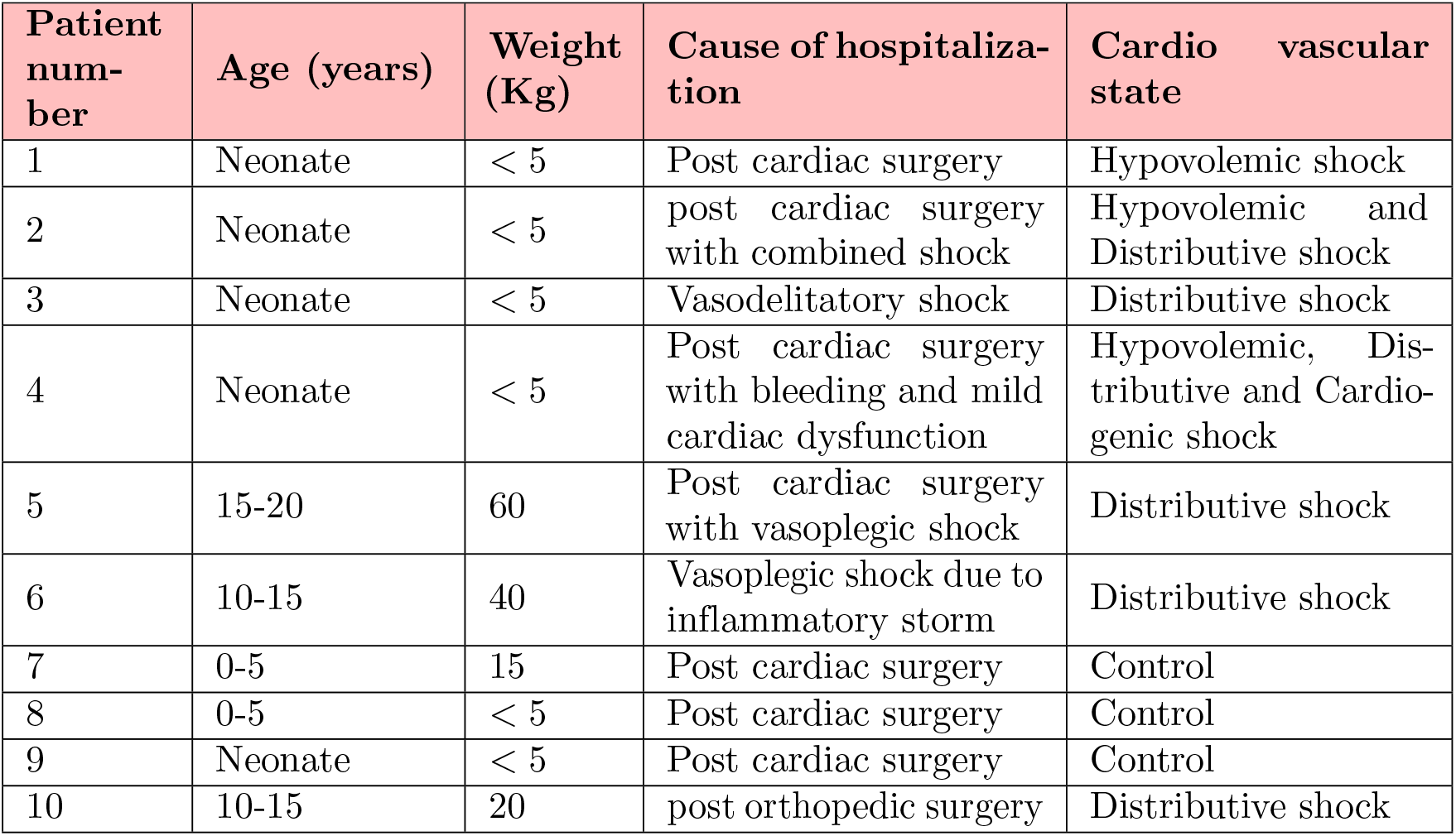
Patients information. For each patient age, weight, cause of hospitalization and cardio-vascular state are given.

| Patient number | Age (years) | Weight (Kg) | Cause of hospitalization | Cardio vascular state |
| --- | --- | --- | --- | --- |
| 1 | Neonate | < 5 | Post cardiac surgery | Hypovolemic shock |
| 2 | Neonate | < 5 | post cardiac surgery with combined shock | Hypovolemic and Distributive shock |
| 3 | Neonate | < 5 | Vasodelitatory shock | Distributive shock |
| 4 | Neonate | < 5 | Post cardiac surgery with bleeding and mild cardiac dysfunction | Hypovolemic, Distributive and Cardiogenic shock |
| 5 | 15-20 | 60 | Post cardiac surgery with vasoplegic shock | Distributive shock |
| 6 | 10-15 | 40 | Vasoplegic shock due to inflammatory storm | Distributive shock |
| 7 | 0-5 | 15 | Post cardiac surgery | Control |
| 8 | 0-5 | < 5 | Post cardiac surgery | Control |
| 9 | Neonate | < 5 | Post cardiac surgery | Control |
| 10 | 10-15 | 20 | post orthopedic surgery | Distributive shock |

**Table 2.** List of measurements.

| Observable | Description |
| --- | --- |
| $\widehat{P_a}$ | Mean arterial pressure |
| $\widehat{P_v}$ | Mean venous pressure |
| $\widehat{P_p}$ | Pulse pressure |
| Hr | Heart rate |
| $\widehat{RC}$ | peripheral resistance multiplied by the arterial compliance and scaled by a constant |

**Table 3.** List of estimated parameters.

| Parameter | Description |
| --- | --- |
| $P_{\text{set}}$ | Set point of the baro reflex |
| $\tilde{C}_v$ | Venous compliance over arterial compliance |
| $\Delta\tilde{V}_{v0}$ | Difference between maximal and minimal unstressed volume over arterial compliance |
| $\tilde{I}_{\text{ex}}$ | Intra vascular volume change over arterial compliance |
| $\text{Hr}_{\text{min}}$ | Minimal heart rate |
| $\text{Hr}_{\text{max}}$ | Maximal heart rate |
| $\tilde{R}_{\text{min}}$ | Minimal peripheral resistance multiplied by arterial compliance and by $\alpha_{RC}$ |
| $\tilde{R}_{\text{max}}$ | Maximal peripheral resistance multiplied by arterial compliance and by $\alpha_{RC}$ |
| $\tilde{K}_{\text{min}}$ | Minimal relative contractility |
| $\tilde{K}_{\text{max}}$ | maximal relative contractility |
| $S_{\text{const}}$ | Constant component of the independent autonomic activation |
| $S_{\text{slope}}$ | Slope of the independent autonomic activation |
| $M_{\text{const}}$ | Constant component of the modulation over peripheral resistance |
| $M_{\text{slope}}$ | Slope of the modulation over peripheral resistance |
| $\alpha_{RC}$ | A constant that scales the peripheral resistance |

As noted above, iCVS is able to deal with patients of varying weights from neonates (a couple of kilograms) to adult-sized patients, as we demonstrate below. The hemodynamic data for all patients in this ICU are collected as routine practice [12], [14]. The diagnostic labels for shock (either hypovolemic/ hemorrhagic, distributive, cardiogenic, or a combination of the above) were derived from a prospective and retrospective review of at least two expert clinicians. These labels were also corroborated using objective measurements from the medical record, when available. For example, from quantification of blood loss postoperatively, assessment of cardiac function or systemic vascular resistance using clinical examination and ancillary diagnostic tests.

The challenge of estimating the causes for cardiovascular instability and shock from real-world data can be appreciated by examining Figs. 5-8. Examination of panels A-E in each figure, which depict the five observables, shows that patients’ data are “messy” with marked fluctuations and trends arising from multiple physiological sources (due to the shock cause and others), unlike the clean simulated data shown before. Moreover, it is evident that simple rules linking changes in these observables to the shock cause may be misleading.

**Fig 5.**
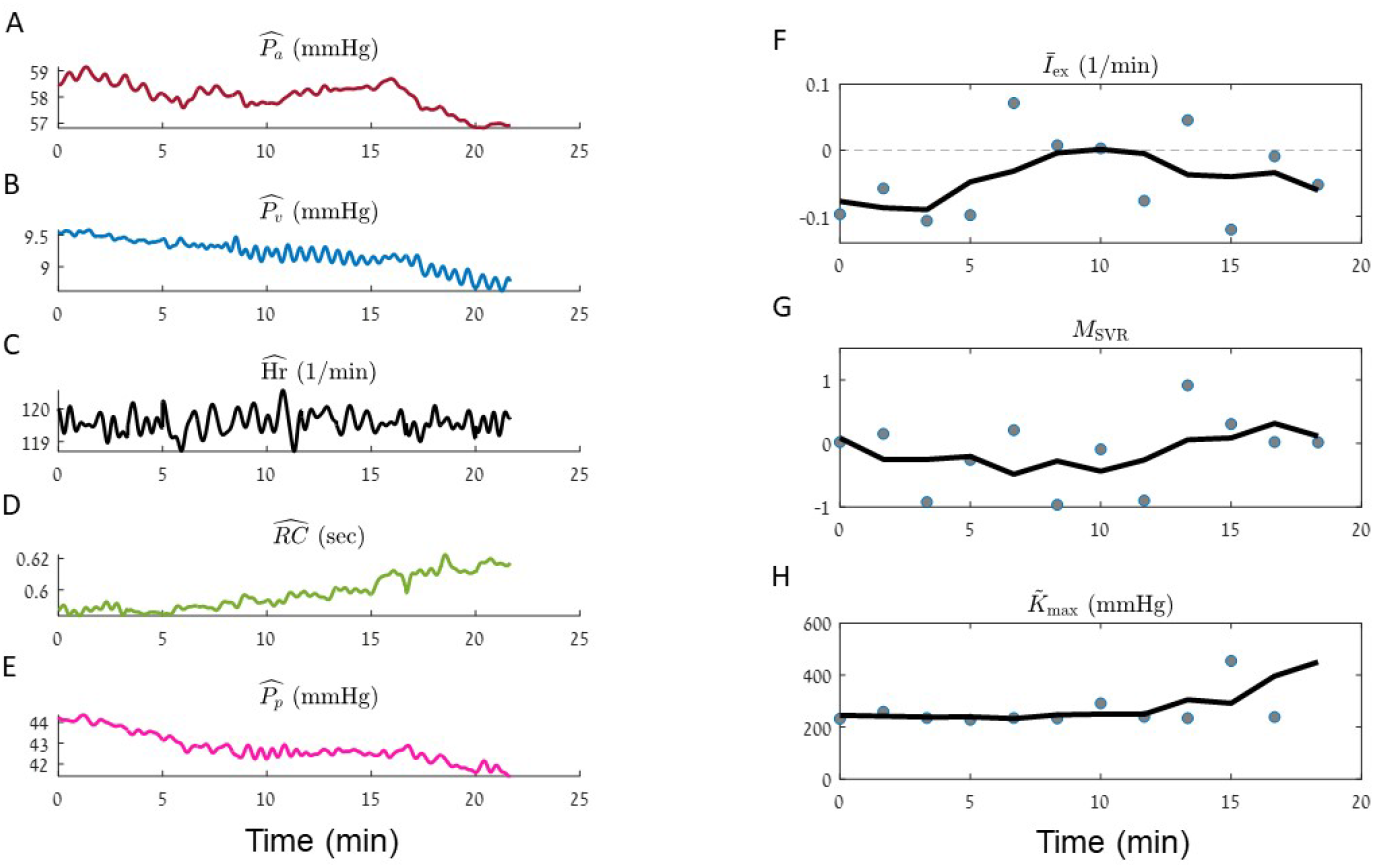
iCVS model inference results for a neonate patient (body weight *<* 5 Kg), in a hypovolemic shock state. iCVS results for patient number 1 (see Table 1) are presented. **A-E**. - The observables: mean arterial pressure (**A**), mean venous pressure (**B**), heart rate (**C**.), RC - the peripheral resistance multiplied by the arterial compliance (**D**) and pulse pressure (**E**). **F-H**. Gray circles - optimal parameters which are achieved in each segment of 300 sec. Black line - a smoothed version of the estimated parameters (see *Material and methods*). **F**. Relative intra vascular volume change (*Ī*_ex_). **G**. Non autonomic vascular resistance (*M*_SVR_). **H**. Maximal relative contractility 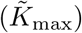. Note that in this patient which suffers from a hypovolemic shock state is identified as having negative *Ī*_ex_ by the model.

**Fig 6.**
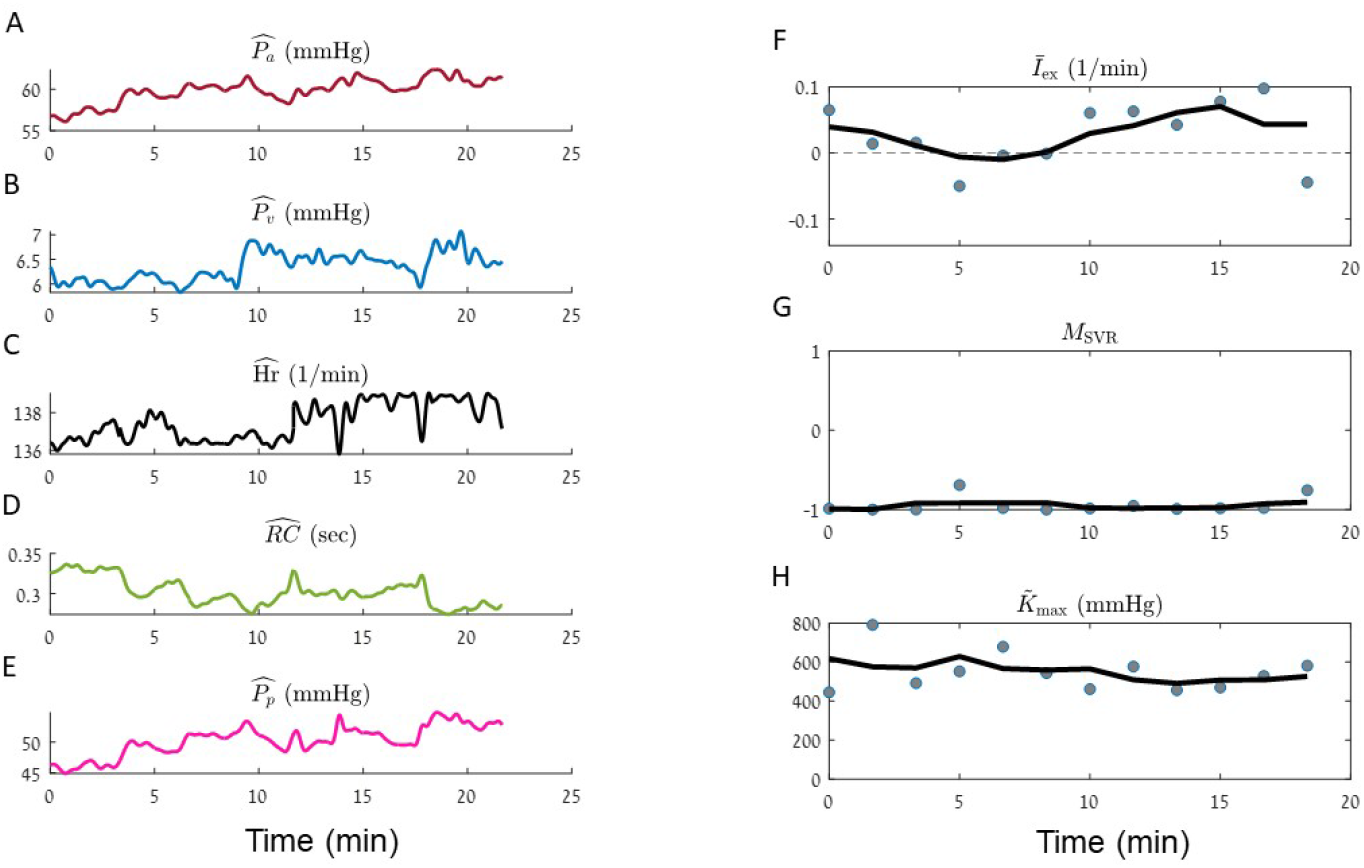
iCVS model inference results for an adolescent patient (body weight 60 Kg), in a distributive shock state. iCVS results for patient number 5 (see Table 1) are presented. **A-E**. The observables: Mean arterial pressure (**A**), mean venous pressure (**B**), heart rate (**C**), RC - the peripheral resistance multiplied by the arterial compliance (**D**) and pulse pressure (**E**). **F-H**. Gray circles - optimal parameters which are achieved in each segment of 300 sec. Black line - a smoothed version of the estimated parameters (see *Material and Methods*). **F**. Relative intra vascular volume change (*Ī*_ex_). **G**. Non autonomic vascular resistance (*M*_SVR_). **H**. Maximal relative contractility 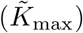. Note that this patient, suffering from a distributive shock state, is correctly identified as having negative *M*_SVR_ by the model.

**Fig 7.**
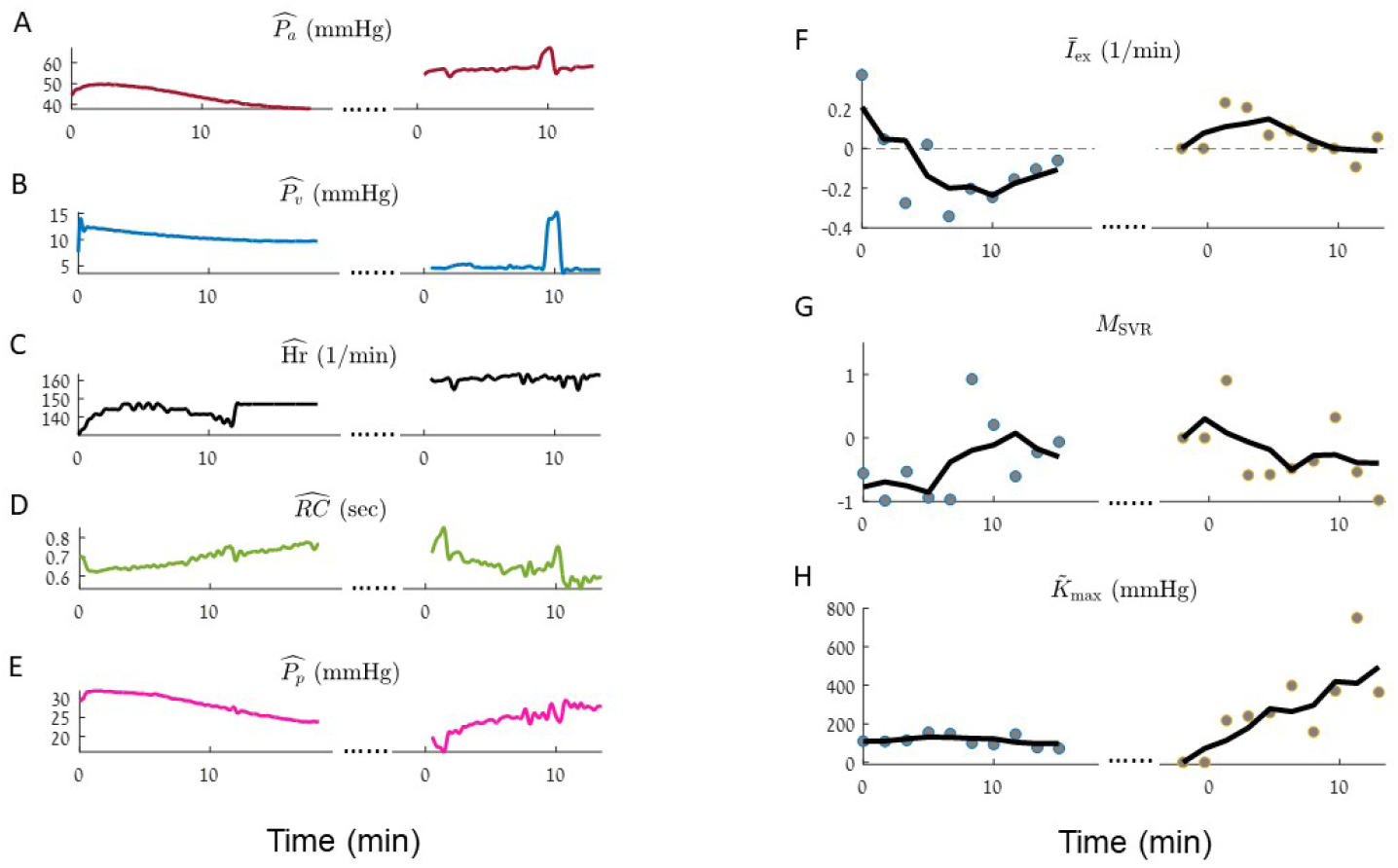
iCVS model inference results for a neonate patient (body weight *<* 5 Kg), in a combined hypovolemic, cardiogenic and distributive shock state. iCVS results for patient number 4 (see Table 1) are presented. Dots mark chest opening. **A-E**. - The observables: mean arterial pressure (**A**), mean venous pressure (**B**), heart rate (**C**), RC - the peripheral resistance multiplied by the arterial compliance and pulse pressure (**E**). **F-H**. Gray circles - optimal parameters which are achieved in each segment of 300 sec. Black line - a smoothed version of the estimated parameters (see *Material and methods*). **F**. Relative intra vascular volume change (*Ī*_ex_). **G**. Non autonomic vascular resistance (*M*_SVR_). **H**. Maximal relative contractility 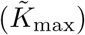. Note that in this patient *Ī*_ex_ becomes non-negative and the contractility increases after the chest opening procedure, compatible with an improving in cardiac function and bleeding cessation as a result of the chest opening procedure.

**Fig 8.**
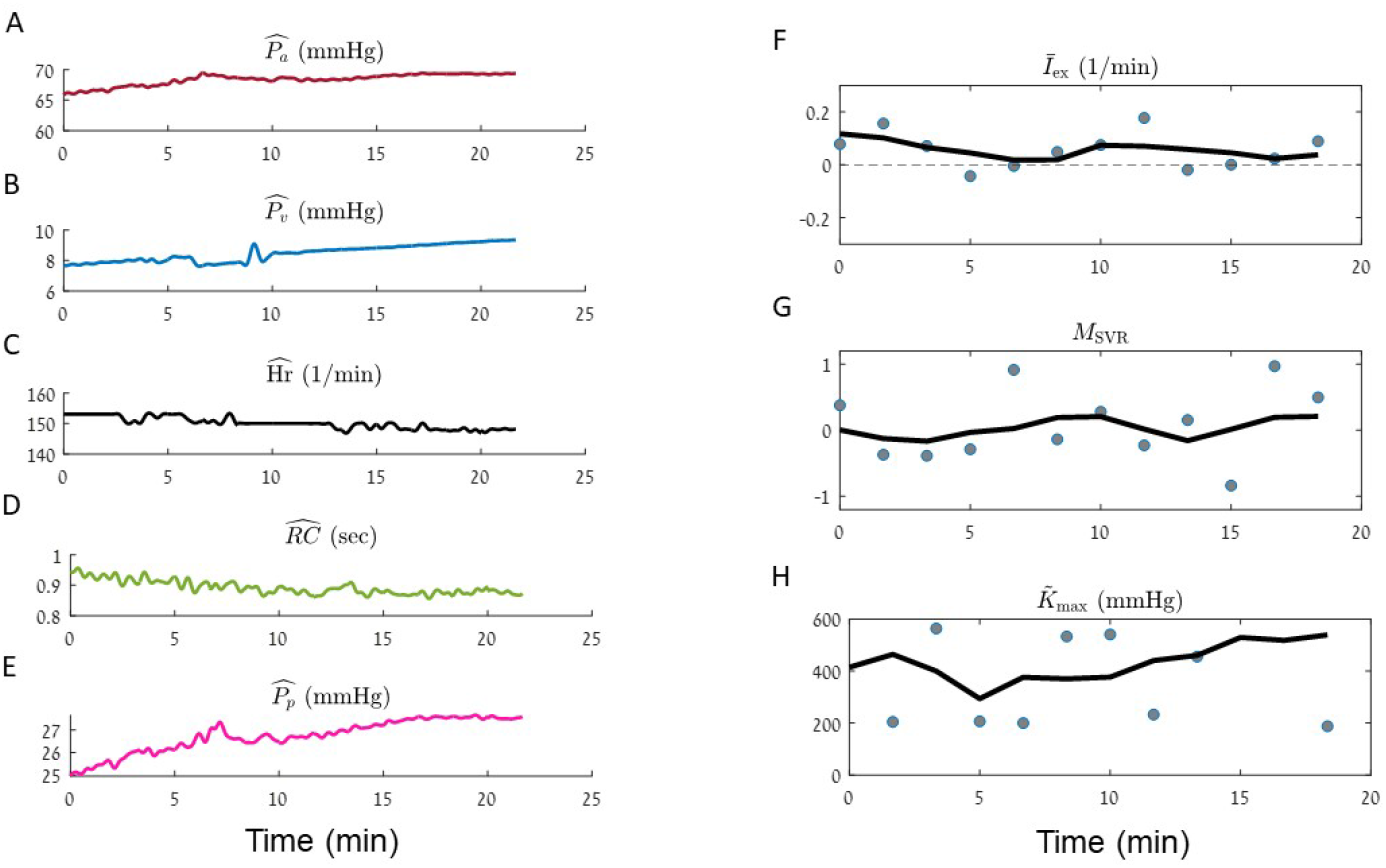
Illustration of the estimation process for a neonate (body weight*<* 5 Kg) control patient, post cardiac surgery. iCVS results for patient number 9 (see Table 1) are presented. **A-E**. The observables: mean arterial pressure (**A**), mean venous pressure (**B**), heart rate (**C**), RC - the peripheral resistance multiplied by the arterial compliance (**D**) and pulse pressure (**E**). **F-H**. Gray circles - optimal parameters which are achieved in each segment of 300 sec. Black line - a smoothed version of the estimated parameters (see *Material and methods*). **F**. Relative intravascular volume change (*Ī*_ex_). **G**. Non autonomic vascular resistance (*M*_SVR_). **H**. Maximal relative contractility 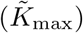. Note that in this control patient *Ī*_ex_ and *M*_SVR_ are not negative and the contractility is stable.

Panels F-H in Figs. 5-8 present the results of model estimation for three hidden parameters that are crucial to elucidate the causes for cardiovascular shock. A hypovolemic state is described by a negative *Ī*_ex_ (the intra-vascular volume change divided by the difference between the maximal and minimal unstressed venous volume Δ*V*_*vo*_). This magnitude does not depend on the patient weight or blood volume, has units of 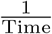, and thus allows comparison between patients with different sizes. The vasodilatatory state is represented by *M*_SVR_ (non-autonomic vascular resistance) which is expected to be negative in a distributive shock state. The cardiac function is described by the *Maximal relative contractility* :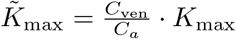, where *K*_max_ is the maximal contractility that the ventricle can generate (see Eq. 21), *C*_ven_ is the compliance of the ventricle, and *C*_*a*_ is the arterial compliance. For constant arterial and ventricular compliances, 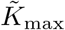 is proportional to the maximal heart contractility. 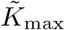 is expected to decrease in case of cardiogenic shock or cardiac dysfunction.

Each gray circle in panels F-H in Figs. 5-8 corresponds to the optimal parameters which are obtained in a given time segment. The black line in each panel is a moving average of the results of different segments (see *Material and methods*).

Fig 5 illustrates model inference for a sample patient with post-operative bleeding. This is a neonate that presented with significant intravascular volume loss after cardiac surgery. Panels A-E show decrease in the arterial and venous blood pressures, while SVR increases and pulse pressure decreases, as expected. Of note is that for an unknown reason, the heart rate of this patient does not reflexively increase, as one would expect for a bleeding patient. Panel F shows that even without the expected heart rate increase, our model correctly identifies the negative intravascular volume change (Fig 5F). In addition, the estimation process yields non-negative *M*_SVR_ (Fig 5G), meaning that the patient is not identified as having a maladaptive vasodilatation. The maximal relative contractility seems stable (for comparison see the changes in 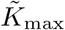 for a patient with cardiac dysfunction in Fig 7).

Fig 6 illustrates the model inference results for an adolescent patient which was clinically identified as having distributive shock, specifically a vasoplegic state seen sometimes after solid organ transplantation. In the time window shown here, the mean venous and arterial pressures increase, as also the heart rate and the pulse pressure. Yet, the mean arterial pressure is low and the heart rate is high, supporting the clinical observation of a shock condition. We can further see that the peripheral resistance decreases over time. Our model indeed correctly identifies the cause for shock: *M*_SVR_ which reflects the non-autonomic component of the vascular resistance is negative, in agreement with the clinically detected distributive shock. Moreover, the estimation process yields roughly zero *Ī*_ex_, meaning that no intra-vascular volume change is detected, as well as demonstrating stable maximal relative contractility.

Fig 7 presents the estimation process for an unstable patient with a combined hemorrhagic, cardiogenic, and distributive shock. This was a neonate with massive bleeding post cardiac surgery, combined with left ventricular dysfunction and inadequate systemic vascular resistance. The patient was extremely unstable despite multiple interventions and underwent a chest opening procedure in the ICU (grey arrows in Fig 7) which resulted in marked improvement in cardiac function and bleeding cessation. We see that arterial blood pressure increases after the chest opening. In addition, the venous pressure decreases - compatible with an improvement in the heart function (Fig 7A,B). Note how the model correctly identifies the marked improvement in the maximal contractility and *Ī*_ex_ immediately after the procedure.

Finally, a neonate control patient that was admitted after thoracic surgery and had an uneventful post-operative course is presented in (Fig 8). As expected, the estimated intravascular volume change (*Ī*_ex_) and the non-autonomic SVR modulation (*M*_SVR_) are not negative, and there aren’t marked fluctuations in the maximal relative contractility 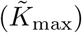.

In Fig 9 we present results summarizing the model’s inference for the 10 patients. Panel A depicts the ability of our model to correctly identify intravascular volume loss: the patients marked in blue are those that had documented post-operative bleeding. Note that for these patients mean change in blood volume is more negative than their counterparts. Conversely, panel B depicts the mean non-autonomic component of systemic vascular resistance modulation. As noted above, a negative value indicates a maladaptive response with vasodilatation, as one can see in distributive shock states (for example sepsis or vasoplegia). The patients marked in red were identified by expert clinicians as having abnormally low SVR. Indeed, overall, these patients were identified by our model as having lower values of *M*_SVR_.

**Fig 9.**
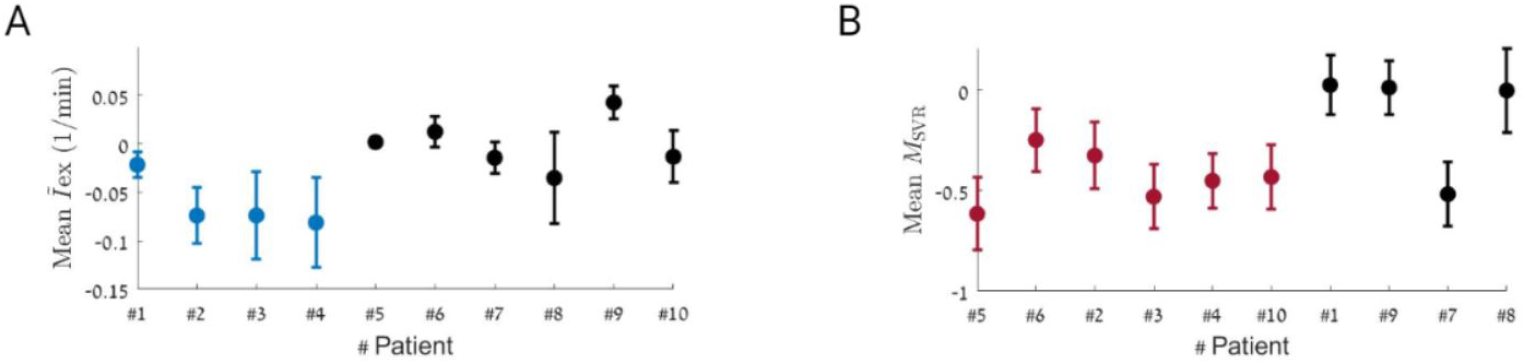
Estimated *Ī*_ex_ and *M*_SVR_ for ten subjects. For each subject, the model is applied for a time window of 25 minutes (see Methods). The analysis is done over segments of 300 seconds and the average values of *Ī*_ex_ (panel **A**) and *M*_SVR_ (panel **B**) are presented. **A**. Average value of *Ī*_ex_ is presented. Blue - patients which are documented with post-operative bleeding, black - no post operative bleeding is documented (average value of the hypovolemic group is −0.06 min^−1^, average value of the non-hypovolemic group is 0.0001 min^−1^, *P*_value_ = 0.041). **B**. Average value of *M*_SVR_ is presented. Red - patients which were identified by an experts as having distributive shock state, black - patients were not documented with distributive shock state (average value of the distributive group is −0.44, average value of the non-distributive group is −0.09, *P*_value_ = 0.077). Error bars in **A** and **B** represent standard error of the mean.

As an additional verification of model estimation performance on real data, we also compare between the measured observables: 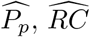 and 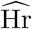 and reconstructed values for the observables which are obtained using model inference. The observables are reconstructed well by the iCVS model, with a Pearson correlation of 0.71-0.94 (see supplementary information). Furthermore, as is elaborated in the in supplementary to this article, we demonstrate the robustness of our findings to the estimation algorithm.

## Discussion

In this work we detail a novel mechanistic model of the cardiovascular system: iCVS. We developed iCVS with a translational goal in mind - enabling estimation of the hidden cardiovascular state of critically-ill patients, in real-time, using only routinely available physiological signals at the bedside. iCVS does not preemptively assume knowledge of almost any parameter, except age and weight, maintaining flexibility for deployment in a variety of clinical scenarios. Unlike many similar models, iCVS considers the autonomic control and its specific components, imposing coupling constraints on inferred parameters and capturing common clinical phenotypes. An additional novel strength of this model is its ability to use slow and fast timescales of the arterial blood pressure waveform to gather more information about the cardiovascular state without increasing model complexity, by performing the main inference on the clinically relevant scale of minutes.

We demonstrate the use of iCVS on a dataset of ten critically-ill children which was collected in a pediatric intensive care unit, illustrating identification of bleeding, distributive states, cardiac dysfunction, and their combination. A key challenge of evaluating models such as iCVS on data acquired from real patients and not in a clean laboratory setting, is the lack of experimentally-defined cardiovascular hidden states. The ground-truth labels for the patients presented in this study were acquired either by information that was available from the electronic health record but which the model was not privy to (e.g. recorded blood loss), or by expert clinician assessment preformed pro- and retrospectively.

While we illustrated the ability of iCVS to identify hidden cardiovascular states that can not be easily identified by simple examination of the physiological recordings, it is obvious that further assessment, ideally prospectively, on a much larger patient cohort is warranted. Generation of a larger dataset, containing a diverse population of patients with various cardiovascular states and their evolution will not only allow model validation and refinement, but also to define cut-off values for shock-state identification. For example, while iCVS can identify ongoing blood loss or maladaptive vasodilatation, ideally we would wish to be able to add an additional layer for a decision support tool that would alert clinicians to the presence of vasoplegic shock or hemorrhagic shock, defined by certain thresholds relating blood pressure to *M*_SVR_ or *Ī*_ex_, respectively. An additional potential limitation of iCVS that can be properly quantified and addressed in future studies is its ability to deal with noisy and artefact-ridden recordings. Moreover, a prospectively collected dataset (that we are currently working towards acquiring) will also allow for precise recording of the various bedside interventions and thus test their effects on the cardiovascular hidden state and model estimation. Finally, despite its high-explanatory power, iCVS is still a relatively simple mechanistic model; it does not take into account various physiological contexts, and therefore cannot address for example the effects of positive pressure ventilation, pericardial effusion or even interactions between the heart chambers and pulmonary and systemic circulations. An additional limitation to be acknowledged is the need for several assumptions in order to enable model simplification and estimation. We attempted to minimize their number and use only ones that are widely accepted, we stated clearly in the methods where such assumptions were made, and also cited their source, where relevant, the literature supporting these assumptions. We also believe, that iCVS can be iteratively developed to include modules that address these limitations, potentially with addition of data-driven machine-learning modules as detailed below.

An exciting avenue for future exploration is to combine iCVS with machine learning models to create robust hybrid clinical decision support systems that can exploit both the power of data-driven analytics and mechanistic models [15, 16]. In the future, we envision such hybrid models deployed at the bedside, providing the clinician with information regarding hidden cardiovascular states and optimal treatment strategies. For example, estimating volume and contractility states, and helping in guiding choices regarding intravenous fluid administration or inotropic support, a current unmet clinical need [17]. The model presented here is the first step towards this ambitious goal.

## Materials and methods

### Mechanistic model

The goal of this section is to detail iCVS: a mechanistic model of the cardio-vascular system that links measurable variables to clinically relevant physiological parameters that can not be measured directly. We use a lumped parameter model with several compartments: the circulatory system is represented as a closed loop with two compliance vessels (the arterial and venous compartments) and one pure resistance vessel (the organs) [5, 18]. The heart contains one chamber that acts as a pump. The venous-arterial flow *I*_*H*_ is generated by the heart and is given by the product of the heart rate Hr and the stroke volume SV:

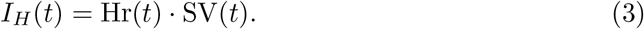

The arteriolar and capillary *I*_*C*_ flows are modeled as a linear resistor:

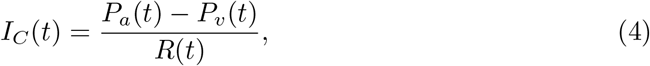

where *P*_*a*_ and *P*_*v*_ are the arterial and venous mean pressures, respectively, and *R* is the peripheral resistance. The evolution of arterial and venous volumes (*V*_*a*_ and *V*_*v*_ respectively) is given by the following differential equations:

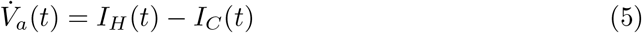

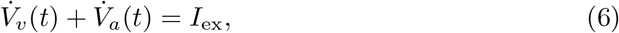

where *I*_ex_ is the intra-vascular volume change - either loss of volume due to bleeding or extra-vascular capillary leak or conversely intravenous fluid administration.

Eqs. 5-6 relate between blood volumes dynamics and other hidden cardio-vascular variables. The challenge however is that blood volumes cannot be directly measured in patients. We therefore reformulate the mechanistic model in terms of blood pressures (which are measurable); this allows us to infer hidden cardio-vascular parameters based on observable data.

The pressure in the venous and arterial compartments is derived from the vessel’s volume and compliance as follows:

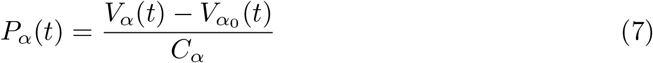

Where *α* ∈ {*a, v*} represents either the venous or arterial components, respectively, *C*_*α*_ the compliance, and 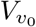 the unstressed volume.

For the arterial compartment, we assume constant unstressed volume [5]. The venous unstressed volume is modulated by an autonomic activation *S*_tot_ (see below):

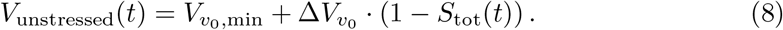

Assuming constant *C*_*a*_ [14], differentiating Eq. 7 and rearranging, we obtain:

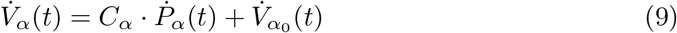

Substitute Eqs. 3, 4 and 9 in Eqs. 5 and 6 we get:

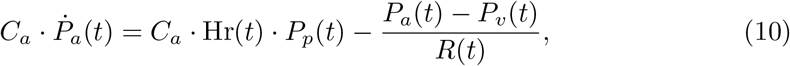

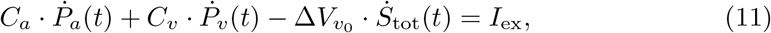

where we assume that the unstressed arterial volume is constant [5] and thus 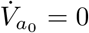, and also use the approximation SV ≌ *C*_*a*_ · *P*_*p*_ [14].

### Autonomic Nervous System control

The model contains elements of autonomic control which affect different physiological quantities: heart rate Hr, peripheral resistance *R*, unstressed venous volume 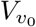 and cardiac contractility *K* (see below). The autonomic control contains two components: the baro-reflex and an external control unit. The baro-reflex, marked by *S*_*b*_, depends on the arterial blood pressure (see also [5]):

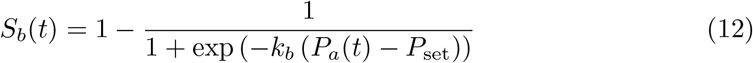

where *k*_*b*_ describes the sensitivity of the baro-reflex and *P*_set_ is the set point of the baro-reflex. In this work *k*_*b*_ = 0.1838 mmHg^−1^ [5].

The autonomic control also contains an additional component, marked by *S* which does not depend on the current arterial pressure. This component is not included in the work of [5]. The overall autonomic activation is a sigmoid function of the summation of the two components:

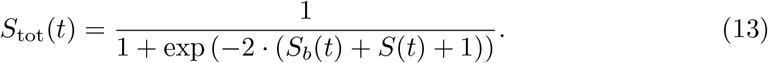

Both *S*_*b*_ and *S* take values between 0 and 1. In addition, the shape of *S*_tot_ is designed such that the total autonomic activation *S*_tot_ is also ranged between 0 and 1, and the value of *S*_tot_ is approximately the mean of *S*_*b*_ and *S*. The two different components of the autonomic activation represent two complementary functions: one is the baro-reflex which is part of a negative feedback loop aimed at maintaining blood pressure and cardiac output homeostasis while the other (*S*) represents the combined effect of sympathetic and parasympathetic activation on the cardiovascular system that is independent of current blood pressure. This division allows us to capture two different behaviours of the cardiovascular dynamics that are commonly observed in critically-ill patients: fluctuations which are negatively correlated between the arterial BP and heart rate, and synchronous fluctuations of the arterial pressure and other components such as heart rate.

#### Heart rate

The heart rate is determined by the autonomic activation, and can vary between minimal and maximal values, Hr_min_ and Hr_max_ respectively:

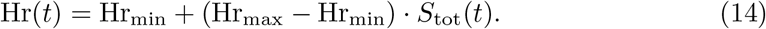

The minimal heart rate (Hr_min_) and maximal heart rate (Hr_max_) are parameters which are approximated by the optimization algorithm with a certain range that depends on the patient age (see Table 4).

**Table 4.**
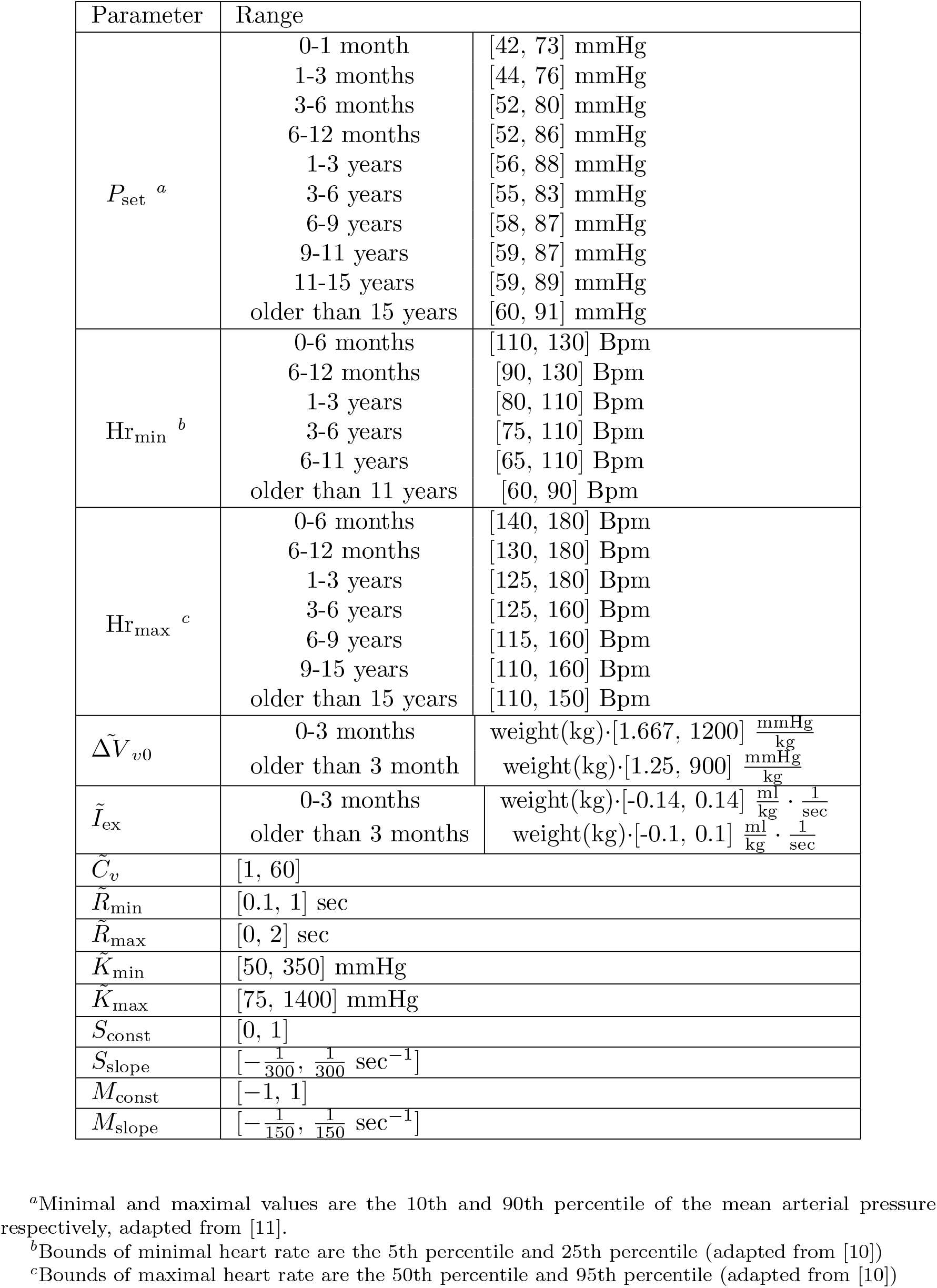
Range of hidden parameters.

#### Peripheral resistance

The peripheral resistance varies between two values, *R*_min_ and *R*_max_, and depends on the autonomic activation and on a local modulation, denoted *M*_SVR_, which does not depend on the autonomic nervous system:

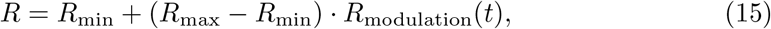

where:

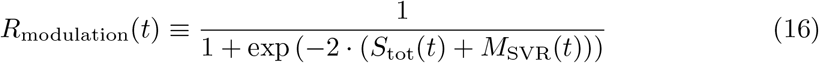

The value of *M*_SVR_ can vary between −1 to 1.

#### Cardiac contractility

The model contains one heart chamber. Of note, Zenker et al. [5], modeled the end systolic volume as a function of the end diastolic volume as follows:

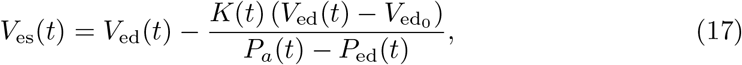

Where *V*_es_ and *V*_ed_ are the end systolic and end diastolic chamber volumes respectively, 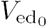 is the unstressed volume of the chamber, *P*_ed_ is the end diastolic pressure of the chamber and *K* is the contractility. Eq. 17 contains information about the heart volumes. Since heart volumes are not directly measurable, we modify the approach adopted by Zenker et al. and derive below an expression that relates heart contractility and blood pressures. We model the heart chamber as a compliance vessel, and estimate the difference between the end diastolic volume (*V*_ed_) and the unstressed ventricular volume as follows:

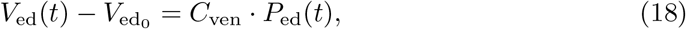

where *C*_ven_ is the compliance of the ventricle. The intra-ventricular pressure (*P*_ed_) cannot be measured directly. Since the heart in the model contains only one chamber, we approximate the end diastolic pressure as the venous pressure:

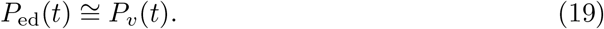

We acknowledge that this approximation is less accurate when the venous pressure is much larger than the pressure during diastole. This can happen for example in case of pulmonary hypertension, valve stenosis, or abnormal heart anatomy. In addition, the stroke volume (SV = *V*_ed_ − *V*_es_), can be approximated using the pulse pressure and the arterial compliance [14]:

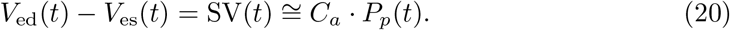

The contractility is modulated by the autonomic activation and is expressed as follows:

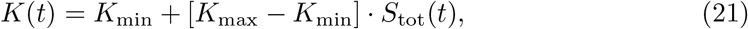

where *K*_min_ and *K*_max_ are the minimal and maximal contractility, respectively. Combining Eq. 17 - 21 we obtain:

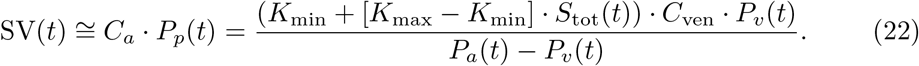

The approximated expression for *C*_*a*_ · *P*_*p*_(*t*) which is presented in Eq 22 can be substituted in *I*_*H*_ which appears in Eq 10 to obtain a system of differential equations for *P*_*a*_, *P*_*v*_.

We have therefore introduced a model that relates between blood pressure dynamics and other cardiovascular parameters. We show below how this mechanistic model can be exploited in order to estimate hidden physiological parameters from routinely bedside collected data.

### Simulations of the model

In Fig 3 and Fig S1 simulations of the iCVS model are presented. The hidden parameters are fixed and the observables are simulated according to Eqs. 10-16, 22. The following values for the hidden parameters are used: *P*_set_ = 50 mmHg, *K*_*b*_ = 0.1838mmHg^−1^, Hr_max_ = 180 Bpm, Hr_min_ = 100 Bpm, *K*_min_ = 100 mmHg, *K*_max_ = 400 mmHg, 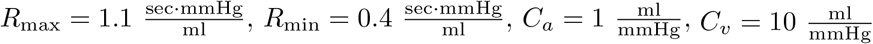, Δ*V*_*v*0_ = 50 ml. In Fig 3 *S* = 0.2. The initial conditions of the mean arterial and venous pressure are set to 48 mmHg and 4 mmHg respectively.

### Extracting the observable variables from the raw data

The raw data which is used in this work is a continuous (sampled at 125 Hz) measurement of the venous and arterial pressure waveforms. The data analysis is performed as follows:

1. Using the algorithm developed by [13, 14], we extract the following measurements from the raw data, all in 10Hz frequency (see Table 2): 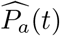(mean arterial pressure), 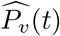(mean venous pressure) 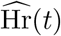(heart rate), 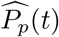(pulse pressure) and 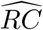, which is an approximation for the peripheral resistance multiplied by the arterial compliance and scaled by a constant (*α*_*RC*_): 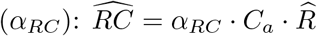.
2. Outliers are found and replaced - Outliers are defined as points which are smaller than the *Lower threshold value* - three local scaled median absolute deviation below the the local median within a sliding window of 20 sec, or higher than the *Higher threshold value* - three local scaled median absolute deviation above the the local median within a sliding window of 20 sec. The outliers are replaced by the following: Lower threshold value for elements smaller than the lower threshold value, and upper threshold value for elements larger than the upper threshold value.
3. A low pass filter is applied and the frequency components above 30 Hz are removed.
4. The data of each subject is divided into segments of 300 sec. Only *Valid* segments are used for further analysis. A valid segment should satisfy the following criteria: 20 mmHg *< P*_*a*_ *<* 250 mmHg, 0 *< P*_*v*_ *<* 25 mmHg, 0 *< RC*, 30 (1*/*min) *<* Hr *<* 250 (1*/*min), 10 mmHg *< P*_*p*_.

### Study approval

The study was approved with waiver of informed consent by the Research Ethics Board at The Hospital for Sick Children, approval number 1000048904.

### Sample subjects analysis

For each subject in Fig 9, the estimated parameters of 12 segments of 5 minutes are averaged. Each segment is 300 seconds long, and the segments are partly overlapped - each segment starts 100 seconds after the previous one. Segments which are not valid are not analyzed.

For the analysis in Fig. 9, for each subject the first recorded 25 minutes time window that satisfies the follows is analyzed: the subject didn’t receive intra-venous fluids bolus during the time window, 15 minuts before and 15 minutes after.

In Figs 5, 6, 7, 8, the above described 25 minutes time windows of patients 1,5,4,9 are presented, respectively.

### The estimation procedure

#### Definition of the cost function

Based on the iCVS model, we define a cost function which consists of a set of terms that link between the observables (Table 2) and the hidden parameters (Table 3). We first define the following expressions: 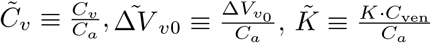 (relative contractility), 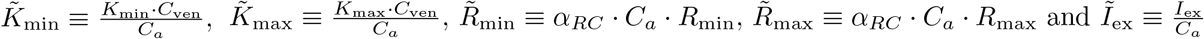.

The cost function is defined as follows:

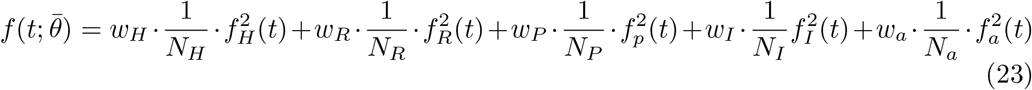

where:

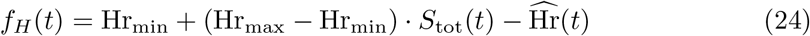

(see Eq 14),

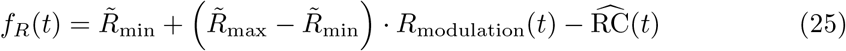

(see 15),

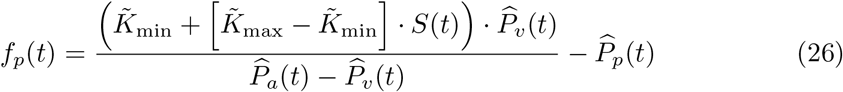

(see Eq 22),

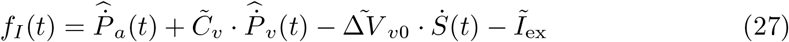

(see Eq 11),

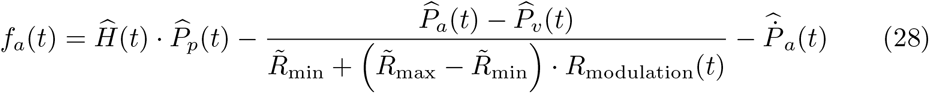

(see Eqs. 10,15).

*R*_modulation_(*t*) is defined in Eq 16, and *S*_*b*_(*t*), *S*(*t*), *M*_SVR_(*t*) are defined below:

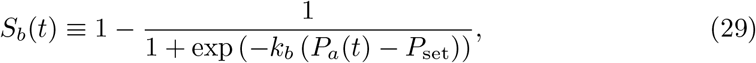

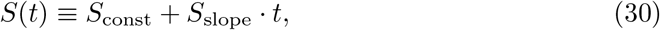

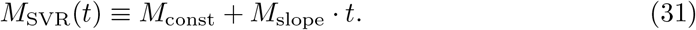

The parameters {*N*_*H*_, *N*_*R*_, *N*_*P*_, *N*_*I*_, *N*_*a*_} in Eq 23 normalize the different terms such that they have no units, and are adjusted in each time interval of the analysis:

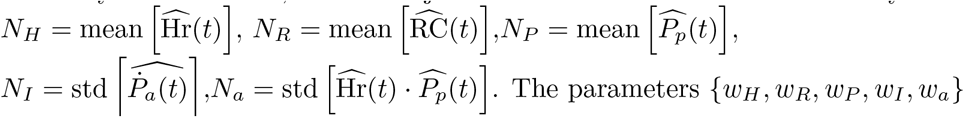

adjust the relative contribution of each term to the cost function. Throughout this work we use the following values: *w*_*H*_ = 1, *w*_*R*_ = 0.2, *w*_*P*_ = 0.2, *w*_*I*_ = 0.2, *w*_*a*_ = 10. The values of *w*_*H*_, *w*_*R*_, *w*_*P*_, *w*_*I*_, *w*_*a*_ are chosen such that the contributions of the terms 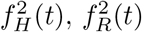 and 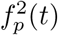 to the cost function have the same order of magnitude, and the contributions of 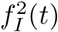 and 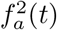 (the terms that contain derivatives) are larger. The optimal parameters are searched within a certain physiological range (see Table 4).

#### Estimation process

Table 3 contains a list of the estimated parameters, where each parameter is bounded within a certain range (see Table 4). The estimation process is done separately in segments which are defined between two time points: [*t, t* + *T*], where *T* = 300 sec. During the estimation procedure, the hidden parameters are estimated by minimizing the time integral over the cost function *f* (*t*) within a given interval. For some of the parameters (see *θ*\* below) we assume that the time dependent change is smooth, and consider also the previous estimated value of the parameters.

The optimization problem can be stated as:

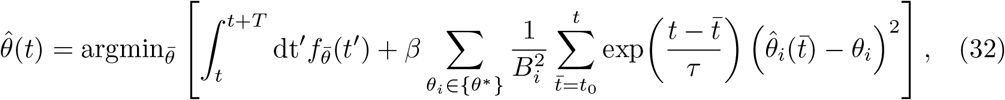

where 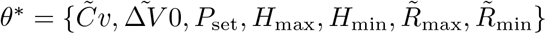 are the parameters for which previous estimator are considered, and *B*_*i*_ is the upper bound of the parameter *θ*_*i*_ (see Table 4). Throughout this work *β* = 0.1.

The optimal solution (the set of optimal parameters 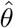) in each segment is chosen using the function *fmincon*.*m* of Matlab with the option *globalsearch*, which searches a global solution for a constrained nonlinear optimization problem across different initial conditions [19]. We supply a random start point to the optimization function.

## Data Availability

All data produced in the present study are available upon reasonable request to the authors

## Acknowledgments

We thank Ori Linial and Ron Teichner for useful discussions. We also thank Robert Greer, Tiffany Ramanathan and Will Dixon for helping with dataset consolidaton. We (US and DE) thank the generous support of ISF 1950/19 and VATAT - council for higher education.

## Supporting information

### S1 Fig. Demonstration of the model for time-varying autonomic control

Fig S1 demonstrates the effect of *S* (independent autonomic control) on the cardio-vascular dynamics in a simulation of the iCVS model. In response to an increase in *S*, arterial pressure, venous pressure, vascular resistance, and pulse pressure increase as well.

**Fig S1.**
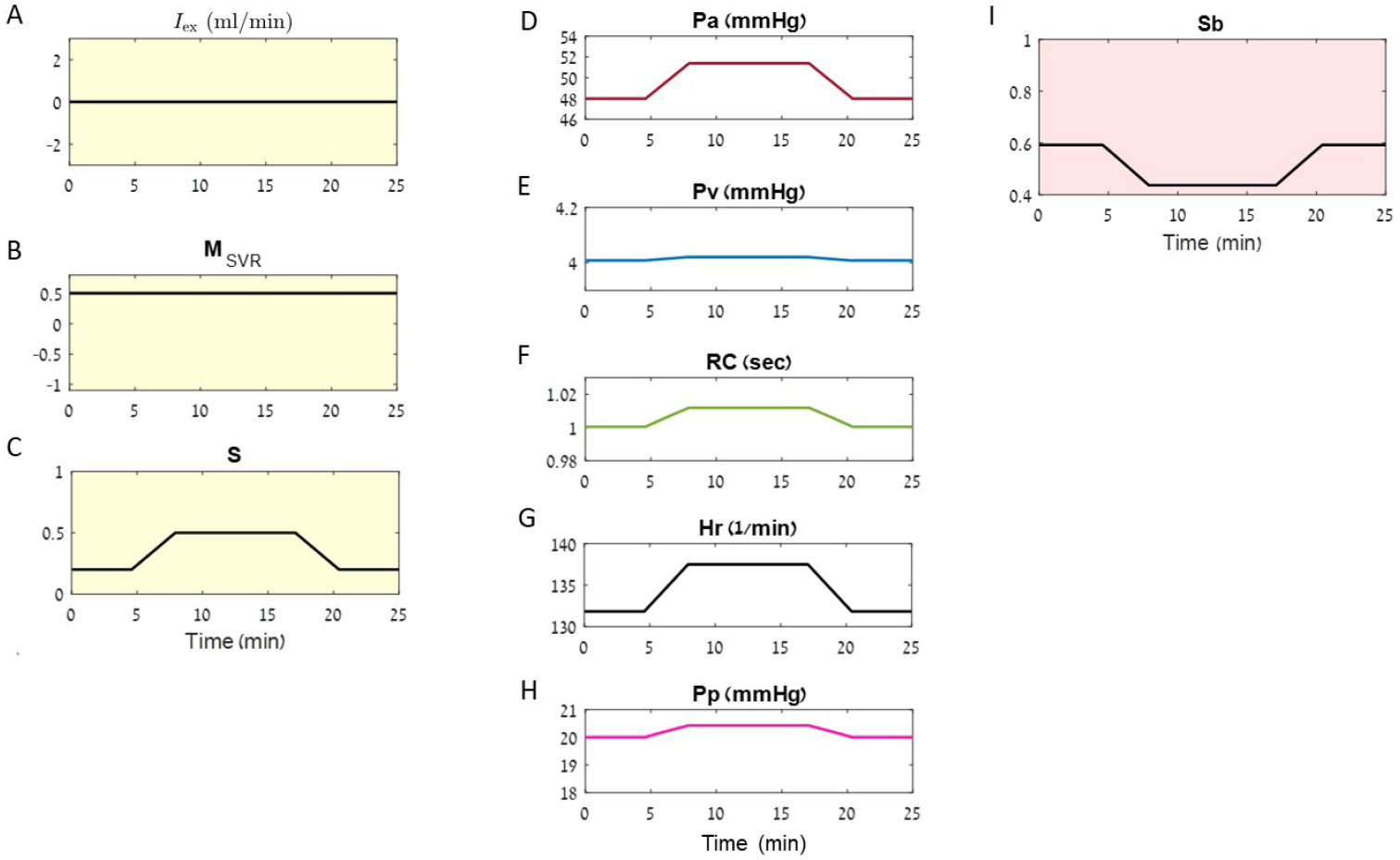
Simulation of the cardio-vascular model - time varying autonomic control. The hidden parameters are fixed and the observables are simulated accordingly (see Methods). **A**. Time dependent intra vascular volume change - zero in this simulation. **B**. Time course of *M*_SVR_ - constant in this simulation. **C**. Time dependent magnitude of the independent autonomic control (*S*) which does not depend on the cardio-vascular state. **D-H** The resulting observables: **D**. Arterial pressure (*P*_*a*_), **E**. Venous pressure (*P*_*v*_), **F**. Peripheral resistance multiplied by arterial compliance (*RC*). **G**. Heart rate (Hr), **H**. Pulse pressure (*P*_*p*_). **I**. Time dependent magnitude of the baro-reflex (*S*_*b*_) which depends on the arterial blood pressure.

**S2 Fig. Reconstruction of the observables** Using the estimated parameters which are found by the optimization process, we reconstruct the heart rate, peripheral resistance and pulse pressure, and compare the reconstructed values to the observed values. The reconstruction is based on Eqs. 24-26, where we define:

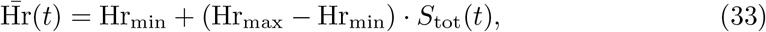

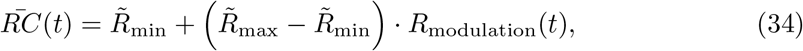

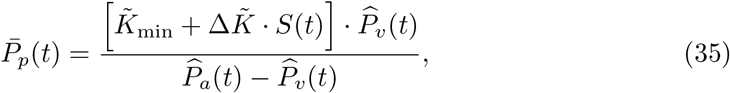

where 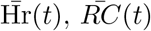 and 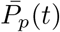 are the reconstructed values of the heart rate, vascular resistance and pulse pressure respectively. Fig. S2 A-C shows examples of reconstructed values vs. observed values. Panels A-C present the reconstruction of the observables for three patients, in a single interval of estimation. Each panel represents a given observable, and each patient is represented by a specific color across panels A-C. Continues line denotes for extracted observables, and dashed line for the reconstructed values. One can see that the iCVS model can reproduce the observables in different scales. The fast fluctuations are not reproduced, becacuse *M*_SVR_ and *S* are modeled as linear in each interval. Fig S2 D-F show the reconstruction for a randomly chosen 1% of the whole data-set which is used in Fig 9. Each dot is a single time point of one patient (all time points of a given patient are represented by the same color). Horizontal axis represents the observed value (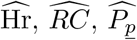 in panels D, E, F respectively), and vertical axis represents the reconstructed value (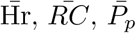 in panels D, E, F respectively). The Pearson correlation coefficient is: 0.917 for Heart rate reconstruction, 0.721 for *RC* reconstruction, and 0.978 for Pulse pressure reconstruction.

**Fig S2.**
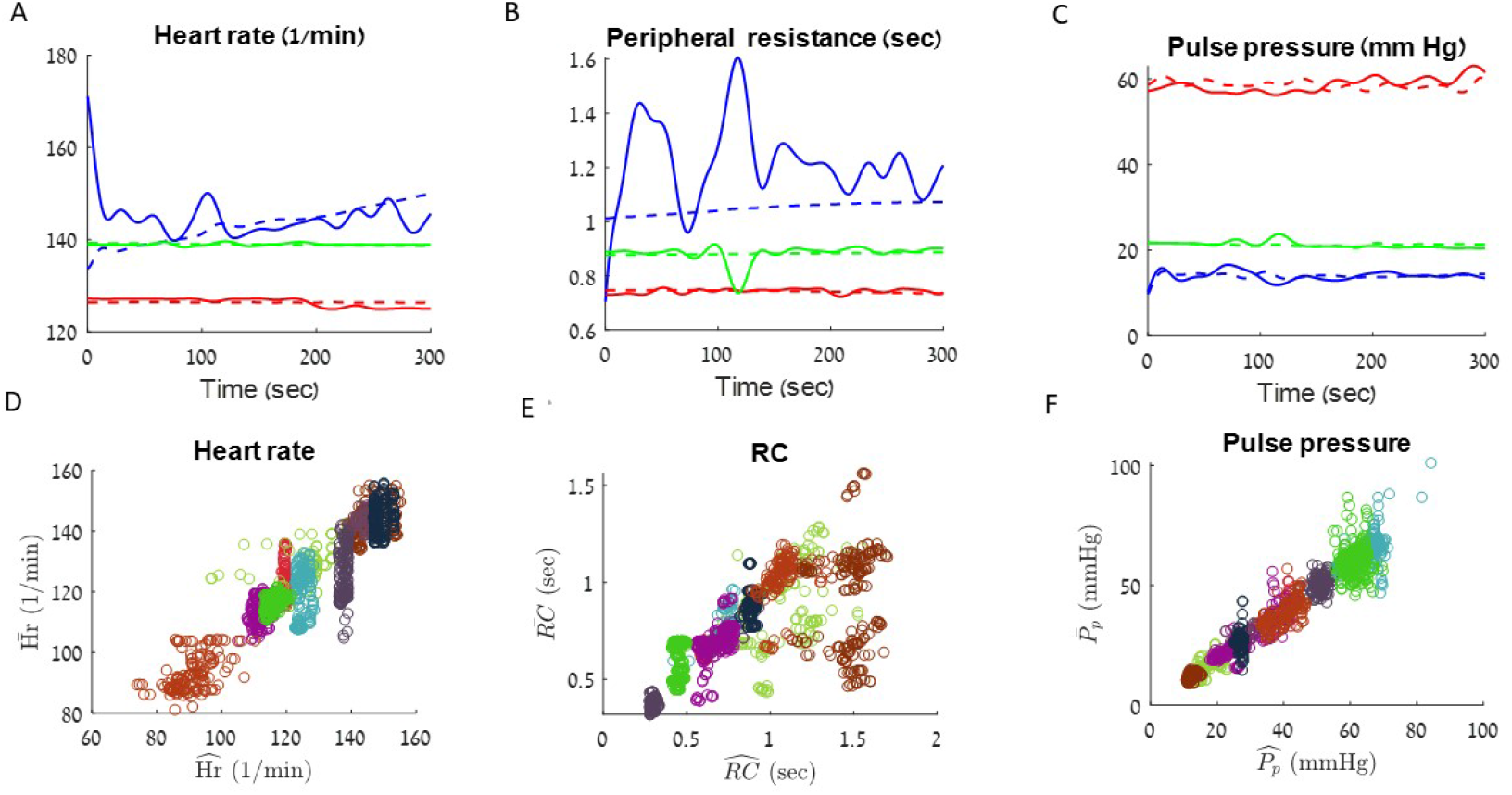
Reconstruction of the observables based on the optimal parameters which are found by the model. **A - C**. Comparison between observed data and reconstructed values during a segment of 500 seconds. Green - patient number 2, blue - patient number 3 and red - patient number 6 (see Table 1). Continuous line - observed data: 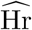(**A**), 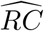(**B**) and 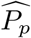(**C**). Dashed line - reconstructed value: 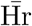(**A**), 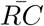(**B**) and 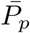. **D-F**. Comparison between the measured value (horizontal axis) and estimated value (vertical axis). Each point represents the value of a given measurement (**D** - heart rate, **E** - peripheral resistance, **F** - pulse pressure) in a specific time point. The time points which are presented are a randomly chosen subset that constitute 1 % of the whole data-set which is used in Fig 9.

**S3 Fig. Robustness of the optimization process**. Fig S3 shows the results of the optimization process for different start points supplied to the optimization function. The illustration is done for patient number 1 which is identified in a hypovolemic shock state and is presented in Fig 5. Panels A-E present the extracted observables. Panels F-H present the optimal parameters obtained for realizations of the optimization process with different start points which are supplied to the optimizaion function. Grey lines denote for different start points, black lines are the results that appear in Fig 5. Purple lines are the average of all the realizations with different start points. It can be seen that the optimization algorithm which is used in this work to fit the iCVS model is affected by the supplied start points. However, when averaging across different initial conditions the results are concordant with the labeled shock state (hypovolemic in this case). In addition, in each single realization the average *I*_ex_ is negative (data is not shown).

**Fig S3.**
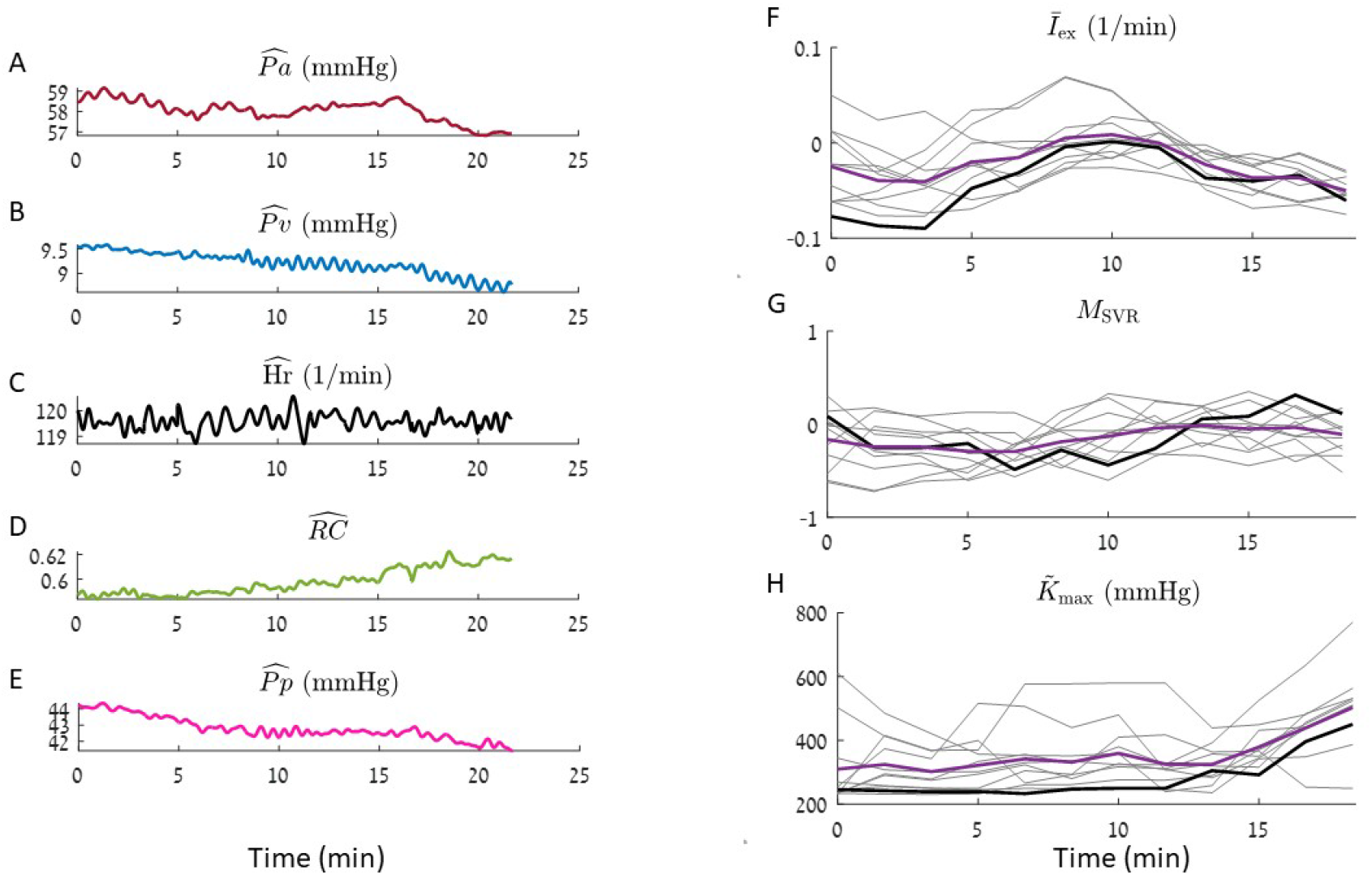
Robustness of the optimization function to different start points. Results for patient number 1 (see Table 1) are presented. **A-E**. - The observables: Mean arterial pressure (**A**), mean venous pressure (**B**), heart rate (**C**.), RC - the peripheral resistance multiplied by the arterial compliance (**D**) and pulse pressure (**E**). **F-H**. Lines represent a smoothed version of the estimated parameters which are obtained in each interval (see Methods). Black line - the results which are presented in Fig 5. Purple line - average over all realizations. **F**. Relative intra vascular volume change (*Ī*_ex_). **G**. Non autonomic vascular resistance (*M*_SVR_). **H**. Maximal relative contractility 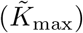.

## Notes

### Competing Interest Statement

The authors have declared no competing interest.

### Funding Statement

This study was funded by grants from the ISF and VATA (the council for higher education in Israel)

### Author Declarations

Ethics committee/IRB of The Hospital of Sick Children (Toronto, Canada) ethical approval for this work

